# Measuring Autonomic Symptoms with the Body Perception Questionnaire Short Form (BPQ-SF): Factor Analysis, Derivation of U.S. Adult Normative Values, and Association with Sensor-Based Physiological Measures

**DOI:** 10.1101/2022.04.27.22274391

**Authors:** Jacek Kolacz, Xiwei Chen, Evan J. Nix, Olivia K. Roath, Logan G. Holmes, Clarissa Tokash, Stephen W. Porges, Gregory F. Lewis

## Abstract

**Objective:** Autonomic regulation of organ and tissues may give rise to disruptions of typical functions. The Body Perception Questionnaire Short Form (BPQ-SF) includes items that were developed to assess autonomic symptoms in daily life. This pair of studies aimed to establish previously unexplored psychometric properties of the BPQ-SF, develop normative values for clinical and research use, and validate the self-reports with sensor-based measures.

**Methods:** Study 1 reports exploratory and confirmatory factor analysis (CFA) on BPQ-SF autonomic reactivity items from a large U.S. population-based online study (n = 2048). In study 2, BPQ-SF scores were examined for associations with heart period, respiratory sinus arrhythmia (RSA), and skin conductance (SC) during seated leg lifts in a community sample (n = 62).

**Results:** Study 1 results supported a 2-factor supra- and sub-diaphragmatic autonomic symptom solution (CFA: *RMSEA* = .040, *CFI* = .99, *TLI* = .99), though a 1-factor solution also fit the data well (*RMSEA* = .080, *CFI* = .99, *TLI* = .99). In study 2, flexible HP responses to lifts and rests were demonstrated at all autonomic symptom levels. However, low self-reported autonomic symptoms were associated with flexible dynamic RSA and SC, moderate symptoms with prolonged SC responses during rest periods, and high symptoms with little systematic changes in RSA and SC during leg lifts.

**Conclusion:** Results support the validity of self-reports of autonomic symptoms in research and clinical applications, with higher symptoms likely indicating impairment in autonomic flexibility.

## Introduction

The autonomic nervous system (ANS), which is responsive to emotional and metabolic challenges, is important for physical health and mental wellbeing. The pathways of the ANS and the brainstem regions with which it interfaces form a rapidly-acting brain-body interface that maintains functional regulation of organs and tissues, integrates information about the state of the body and environment, and controls responses to perceived and anticipated challenges and threats. (1–6) This system provides a physiological component of emotional arousal and regulation, (7–9) and is functionally integrated with brain regions that are involved in safety and threat appraisal (10) as well as the muscles of the face and head that are involved with social signaling of emotions. (11, 12) Its role in coordinating brain and body makes the ANS important for mental and physical health, with disrupted autonomic function being documented in PTSD, anxiety, autism, disorders of the gut, and chronic pain conditions such as fibromyalgia. (13–18) Furthermore, experimental evidence shows that alterations in caregiving environments can promote long-term changes in autonomic regulation (19–21), demonstrating how regulation of autonomic state is sensitive to social tuning and supporting the use of ANS state as an intervention target. The research and clinical importance of the ANS has led to a proliferation of sensor-based tools that quantify its function. (22)

A complimentary and scalable source of ANS information can be acquired using self-reports, which document individuals’ experiences of the body’s function. Self-reports have valuable features as part of an assessment battery. First, self-reports are free or inexpensive to administer, and easy to implement at large scale for research, clinical monitoring, telehealth, and home use. In addition, measures of self-reported autonomic symptoms can contribute information about a range of ANS-innervated targets, compared to sensor-based measures which typically provide targeted information that quantifies performance of a single target organ. Although the function or dysfunction of any one organ can have a unique cause [e.g., tissue damage or a precise response to an environmental condition; (5)], combining information across multiple ANS-innervated targets provides a profile of underlying autonomic regulation. Despite these benefits, self-reports with established psychometric properties and validation are lacking.

### The Body Perception Questionnaire (BPQ-SF)

The BPQ-SF (23; 24) is a self-report questionnaire developed to quantify experiences of body functions. The questionnaire includes autonomic reactivity subscales, which measure the disruption of organs innervated by the ANS. Items indexing autonomic reactivity are informed by the polyvagal theory (11,12,25), which posits that the brainstem structures regulating the ANS are integrated with the neural regulation of muscles of the face, neck, and head through the ventral vagal complex (VVC). Efferent pathways from the VVC innervate the heart, bronchi, esophagus, salivary, and lachrymal glands via visceromotor pathways, which are classically labeled as *autonomic* following Langley’s taxonomy. (1) However, outflow from the nucleus ambiguus of the VVC also innervates the laryngeal and pharyngeal muscles that are involved in swallowing and vocalization via somatomotor pathways, in addition to other muscles of the neck and face. (11) Informed by this neuroanatomical model of brainstem integration centers, the autonomic reactivity items of the BPQ include functions that are coordinated via visceromotor pathways (e.g., “I feel shortness of breath”) and those that include somatomotor pathways (e.g., “I have difficulty coordinating breathing and talking”).

#### Dimensionality

Data from 3 independent samples have supported two autonomic reactivity BPQ factors, corresponding to supra- and sub-diaphragmatic symptoms (above and below the diaphragm; Figure 1), with higher scores indicating greater autonomic disruption of typical organ function (24). This factor structure parallels the innervation patterns of the vagus nerve, which has outflow to structures above the diaphragm that emerge from the nucleus ambiguus (NA) and below the diaphragm from the dorsal motor nucleus of the vagus (DMNX; though both innervate the heart and bronchi). Subsequent studies with Italian and Chinese translations replicated this general factor structure. (26–28)

**Figure 1.**
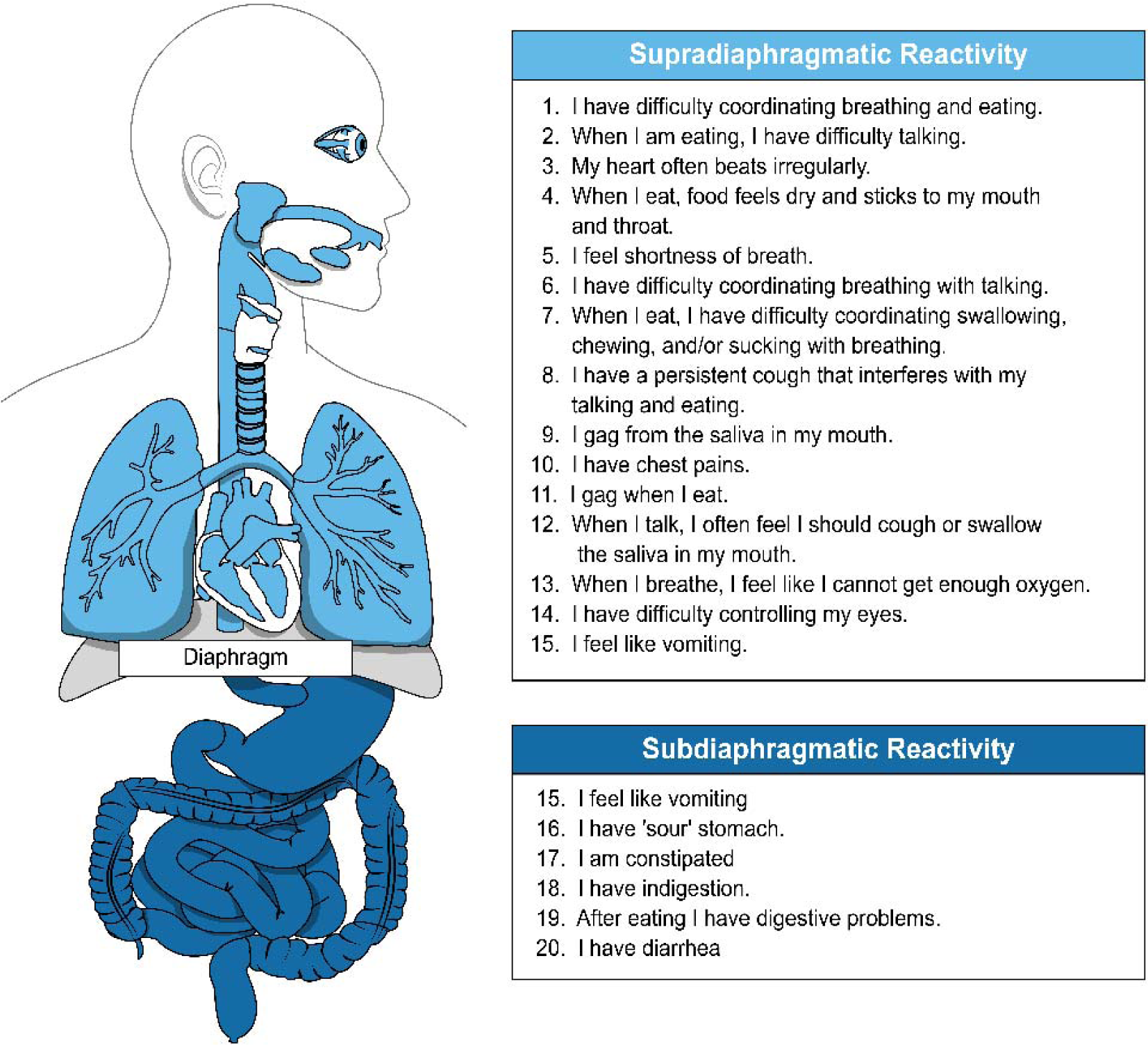
The Body Perception Questionnaire-Short Form (BPQ-SF) autonomic reactivity subscales are composed of items that measure experience of disruptions in circuits that affect function above and below the diaphragm.

Because the BPQ has an ordered categorical response scale (5-point Likert-type scale ranging from “never” to “always”), analysis of item level data requires ordinal methods to avoid statistical distortions. (29) Sample size limitations of initial studies and subsequent factor analysis replications have resulted in the full ordinal outcomes becoming truncated to binary or 3-level responses to avoid overfitting the data influence of random noise. (24,26,27) These strategies have resulted in replicable, interpretable factor solutions but it is unclear whether the factor structure applies to full-item distributions, which are commonly used for subscale scoring. Thus, factor analysis on the English BPQ-SF with the full item distributions is needed.

#### Norms

Prior English-language studies have relied on convenience samples, limiting the generalizability of resulting score descriptive statistics. Normed values from a population-based sample are needed to inform score interpretation and clinical application.

#### Validity

The use of the BPQ to measure body awareness has been detailed in other studies, showing that BPQ scores are correlated with activity in brain regions associated with interoception (30) and research has suggested that the awareness subscale is a measure of interoceptive sensibility (31) or awareness of neutral and uncomfortable body sensations. (32) Associations of the body awareness subscale and other similar measures are explored in other studies. (24,27,32,33) However, to our knowledge, there has up to now been no systematic research to explore the association of the BPQ ANS reactivity subscales with sensor-based measures of physiological activity.

### The Present Studies

Study 1 utilized a large national U.S. sample to examine the full-item response factor structure of the English-language Body Perception Questionnaire Short Form (BPQ-SF) and applied the results to develop population-based percentile and T scores. We hypothesized that factor analysis would converge on a structure similar to that which emerged from binary or three-level simplification of item response scales in past studies.

Study 2 examined the associations of BPQ-SF autonomic reactivity subscales with sensor-based autonomic measures during seated leg raises. Electrocardiogram (ECG) and electrodermal activity (EDA) were used to assess: 1) changes in mean heart period, a broad metric of cardiac output, 2) respiratory sinus arrhythmia (RSA) amplitude, also known as high frequency heart rate variability, a measure of myelinated parasympathetic regulation, (34) 3) skin conductance reactivity frequency [an index of sympathetic regulation; (35)], and 4) vagal efficiency, a measure of the coordination of simultaneous changes in parasympathetic activity and heart period. (36, 37)

According to principles of allostasis, (6, 38) flexible regulation of physiological activity to metabolic demands are key to maintaining adaptive responding, while less dynamic capability place an individual at risk of experiences of disrupted function. Given these principles, we hypothesized that higher BPQ autonomic reactivity scores would be related to reduced dynamic range of the ANS across sympathetic and parasympathetic metrics. We expected that high self-reported symptoms would be associated with less metabolic change in response to leg lifts, poorer ability to return to baseline during recovery periods (legs down), and lower vagal efficiency across the task period.

Study 1.

## Method

### Procedure

The study was approved by the Indiana University Institutional Review Board. All participants provided informed consent. Data were collected using an online survey consisting of the BPQ-SF, self-reported demographics, and several questionnaires not related to the present study. U.S. residents age 18 years and older were recruited through Qualtrics’ research panels, selected based on age, race, and gender to reflect the U.S. general population. Online panel data have been found to have similar internal reliability estimates and effect sizes between variables compared to conventional sampling methods (39) and U.S. residents recruited through Qualtrics are demographically similar to a national probability sample. (40) Recruitment and data quality check details are in Supplemental Materials.

Factor analysis. Dimensionality was assessed by exploratory and confirmatory factor analysis (EFA and CFA) using the packages “psych” (41) and “lavaan” in R 4.1.1. (42) Factor analyses were applied to full item distributions using polychoric correlations and a weighted least squares estimator. (43) Exploratory factor retention was guided by loading structure, parallel analysis, and model fit. (44) Parallel analysis was conducted to examine the number of eigenvalues that deviated from the randomly simulated and re-sampled data, (45) with factors within +/-1 of the deviation from random data considered as viable solutions. (46) Goodness of fit was evaluated using the root mean squared error of approximation [RMSEA; (47, 48)], the Tucker-Lewis Index [TLI; (49)]; and the Comparative Fit Index [CFI; (50)]. Adequate fit was evidenced by an RMSEA value near .06 or lower as well as CFI and TLI values near .95 or greater. (51) EFA results were subject to geomin oblique rotation, (52; 53) which can reproduce correlated and uncorrelated factor structures. (44) The EFA solution was then applied to the second subsample using CFA and assessed using the fit indices described above.

Calculation of percentiles and T scores. Percentiles were calculated from sum scores of subscales based on the factor analysis result. T-Scores were derived by normalizing ranks to a mean of 50 and standard deviation of 10. Unobserved scores were interpolated with a monotonic Hermite spline function. (54)

## Results

The study 1 cohort was composed of 2048 adults. Descriptive statistics are in Table 1. The mean age was 46.34 years (SD = 17.19, range: 18-95) and 50.6% were female. The racial and ethnic distribution of the sample was similar to the general population. (55) Education level was diverse (14.9% graduate or professional degree, 66.9% at least a college degree, and 28.8% high school diploma). Responses were randomized into exploratory and confirmatory subsamples (each n = 1024).

**Table 1.** Sample descriptive statistics for Study 1. Continuous variables are summarized with means and standard deviations. Categorical variables are summarized by counts and percentages.

|  | **Descriptive**<br>**Statistics** |
| --- | --- |
| n | 2048 |
| Age | 46.34 (17.19) |
| Gender (%) |  |
| Female | 1032 (50.6%) |
| Male | 989 (48.5%) |
| Genderqueer | 6 (0.3%) |
| Transgender Man | 8 (0.4%) |
| Transgender Woman | 4 (0.2%) |
| Race and Ethnicity (%) |  |
| White, Caucasian, or European | 1182 (58.7%) |
| Black or African-American | 319 (15.8%) |
| Hispanic or Latino/a | 254 (12.6%) |
| Multiracial | 125 (6.2%) |
| Asian | 102 (5.1%) |
| North American Native | 32 (1.6%) |
| Education (%) |  |
| Less than high school graduate | 64 (4.2%) |
| High school graduate | 436 (28.8%) |
| Vocational or technical degree | 103 (6.8%) |
| Associate degree | 225 (14.9%) |
| Bachelor's degree | 458 (30.3%) |
| Graduate or professional degree | 226 (14.9%) |
| Autonomic Reactivity (BPQ-SF) |  |
| Supra-diaphragmatic Reactivity |  |
| Raw Score | 23.63 (11.17) |
| T Score | 48.92 (10.28) |
| Sub-diaphragmatic Reactivity |  |
| Raw Score | 10.99 (4.77) |
| T Score | 49.33 (9.10) |
| Total Autonomic Symptoms |  |
| Raw Score | 34.62 (15.11) |
| T Score | 48.91 (10.16) |
| BPQ-SF = Body Perception Questionnaire Short Form |  |

Exploratory factor analysis. Parallel analysis and fit indices suggested 2-5 factors. Examination of loadings showed that solutions with greater than 2 autonomic reactivity factors violated simple structure, with multiple factors having strong loadings on the same items and some items lacking any substantial loadings (Supplemental Materials). Thus, the 2-factor solution, corresponding to supra- and sub-diaphragmatic reactivity, was retained for CFA (Table 2). There was moderately high correlation between the two factors (r = .79).

**Table 2.**
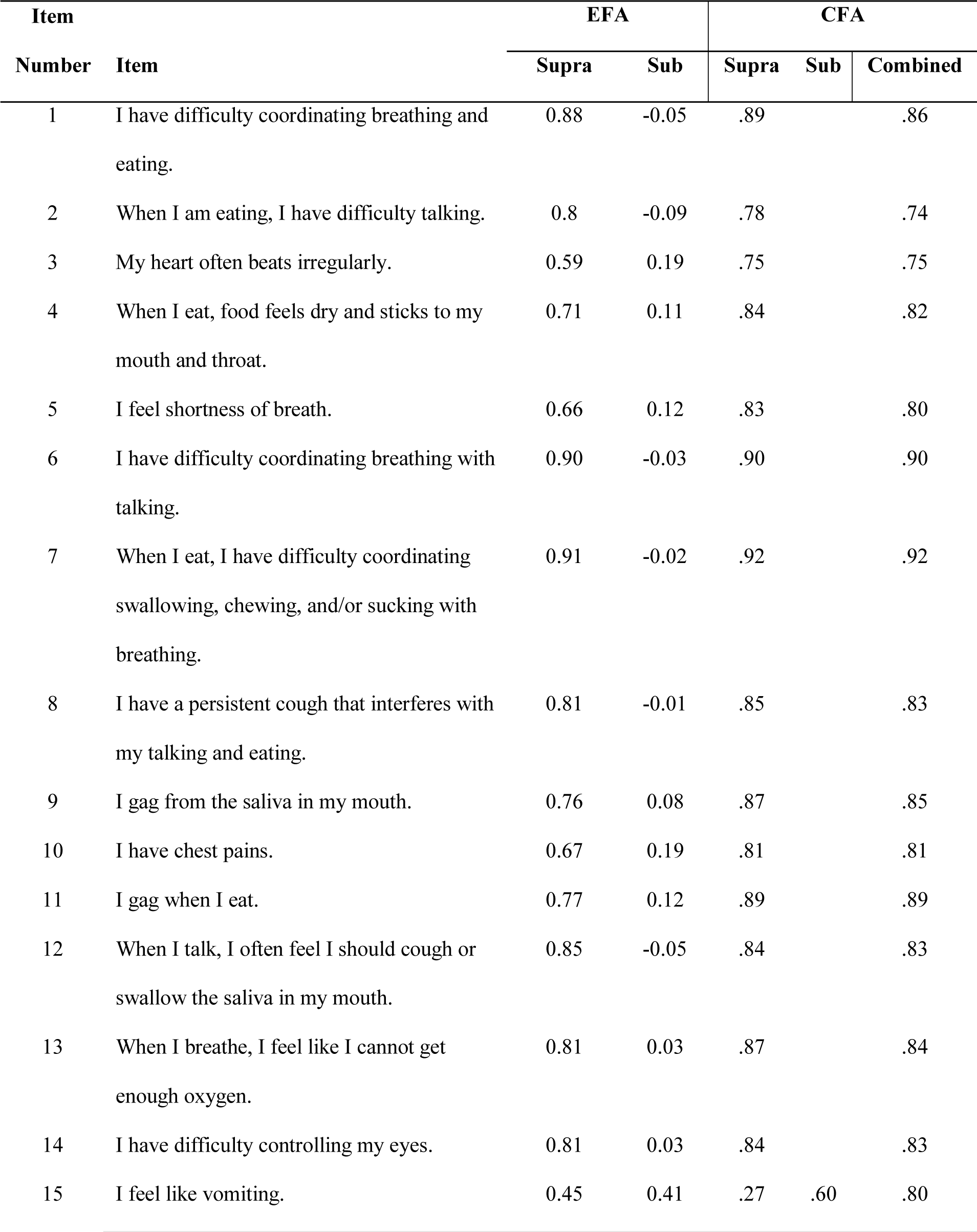
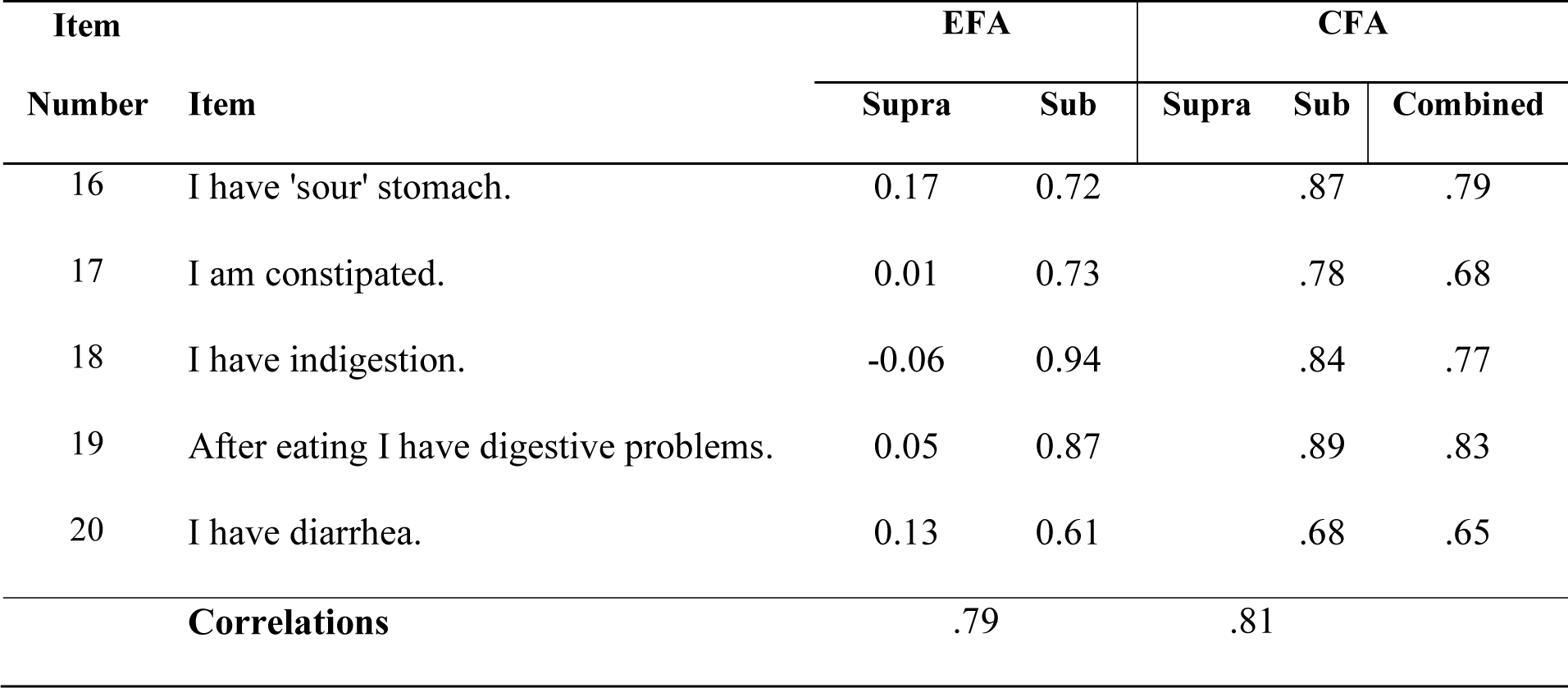
Standardized exploratory and confirmatory factor analysis (EFA and CFA) loadings for the autonomic reactivity subscales of the Body Perception Questionnaire Short Form (BPQ-SF).

Confirmatory factor analysis. The 2-factor solution fit well (*RMSEA* = .044 [90% CI: .039, .049], *CFI* = .99, *TLI* = .99). CFA loadings were similar to those found in the EFA (Table 2). Due to high correlation between factors (*r* = .81), we tested a 1-factor autonomic reactivity solution, which approached good fit (RMSEA = .081 [90% CI: .076, .085], CFI = .99, TLI = .99).

Derivation of Percentiles and T Scores. Tables of percentile rank and T scores are in Supplemental Materials. Correlation between supra- and sub-diaphragmatic T scores was high (*r* = .73).

Study 2.

## Method

### Procedure

The study was approved by the Indiana University Institutional Review Board. Participants were recruited via flyers and screened for eligibility by phone and email. Exclusion criteria were: (1) medication use for heart conditions or blood pressure, (2) a pacemaker, (3) hospitalization for a head injury or major surgery in the past year, and (4) skin allergies.

All participants provided informed consent and completed a demographic survey and the BPQ-SF on a laptop. They were then prepared for physiological data recording of electrocardiogram (ECG) with Lead II configuration and electrodermal activity (EDA) with electrodes applied thenar and hypothenar eminences of the right palm. Data were recorded at 1 kHz using AcqKnowledge Software 4.2 and the Biopac MP160 Data Acquisition System (BIOPAC Systems, Inc., Camino Goleta, CA, USA). For equipment and protocol details see Supplemental Materials.

Seated straight-leg raises (SLR) were used to stimulate an autonomic response. This task was selected for its ease of execution, reliable demand on metabolic load, and low probability of leg movement disturbing sensors on the upper body and hands. First, participants sat quietly for a 2-minute baseline. Participants were then cued through 3 leg lifts during which they extended their legs for 30 seconds with shins parallel to the floor, followed by a 60-second rest with feet down (Figure 2, top panel). Research assistants marked timings at which legs were completely lifted and lowered. All participants were able to keep legs elevated for the full length of all 3 lifts.

**Figure 2.**
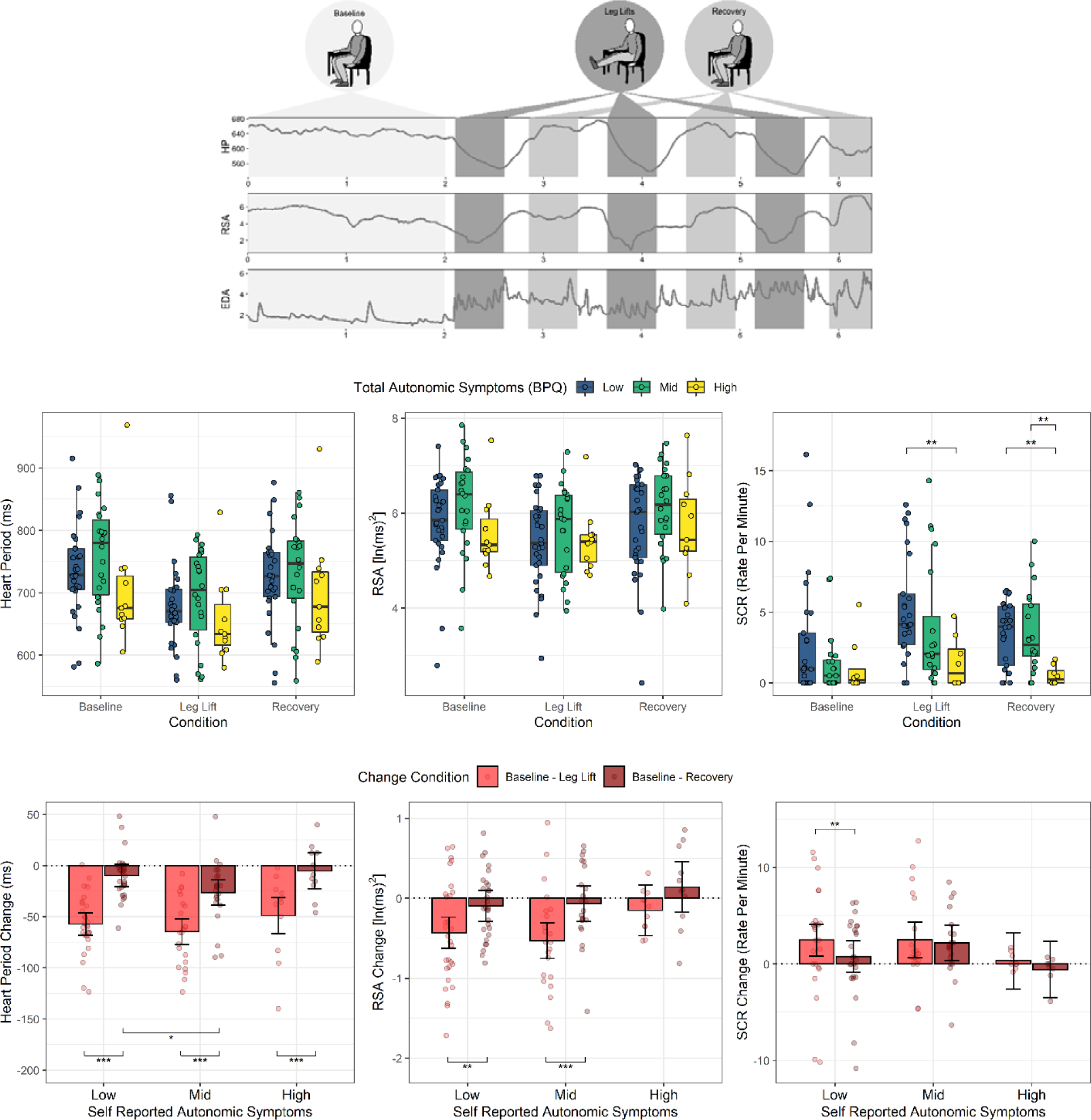
Autonomic activity changes during seated leg lifts as a function of level of autonomic symptoms self-reported on the Body Perception Questionnaire Short Form (BPQ-SF). Top row: Mean heart period (HP), respiratory sinus arrhythmia (RSA), and skin conductance reactivity (SCR) rate were measured during seated baseline, leg lifts, and recovery (legs on floor). Middle row: Raw data and boxplots with between-group significance tests from repeated measures models of absolute values at baseline, leg lifts, and recovery periods. For within-group contrasts see Supplemental Materials. HP and RSA models are adjusted for age and BMI. Bottom row: Linear mixed model predicted values and 95% confidence intervals for HP, RSA, and SCR rate change scores from baseline to leg lifts, and from baseline to recovery periods as a function of self-reported autonomic symptoms. Dotted horizontal lines designate no change.

### Data cleaning and processing

Physiological data cleaning and processing was conducted using AcqKnowledge 4.2 (BIOPAC Systems, Inc., Camino Goleta, CA, USA) and Matlab R2020a. (56) Inter-beat intervals (IBI) were measured by the timing between R-wave peaks from ECG data. IBIs were manually inspected for artifacts, arrhythmias, and missed beats, which may lead to bias and invalidation of beat-to-beat cardiac measures. (57) Missed beats were reconstructed, noise artifacts corrected, and ventricular arrhythmia fluctuations were removed to produce a clean, uninterrupted beat-to-beat signal using CardioEdit+ Software (Brain-Body Center for Psychophysiology and Engineering, University of North Carolina at Chapel Hill). Cleaned data were analyzed in 15-sec epochs. The first and last epochs of recovery periods were excluded from analysis to avoid transition periods and anticipatory autonomic activation immediately before and after leg lifts.

#### Heart period (HP)

HP was quantified as the mean time between successive heart beats in milliseconds, reflecting the sum of all influences on heart beat timing including sympathetic and parasympathetic effects.

#### Respiratory sinus arrhythmia (RSA) amplitude

RSA amplitude is an index of cardiac vagal tone, the effect of the outflow of the nucleus ambiguus via the myelinated vagus nerve pathway to the heart. (34) Calculations were conducted with the Porges-Bohrer method, (58, 59) using a 51-point moving polynomial filter and a second finite impulse response (FIR) type bandpass filter adapted for a 5-Hz resampling rate of the edited IBI time-series. Variability amplitude was quantified in the .12 - 1.0 Hz band, based on the respiratory frequency associated with spontaneous breathing in adults (.12 - .40 Hz) and expanded to include potentially faster respiration rates during the leg-up condition. (34) The result was natural log transformed to reduce skew.

#### Vagal efficiency (VE)

VE indexes the extent to which changes in cardiac vagal tone are associated with changes in cardiac output (36,37,60), calculated from within-individual slopes of synchronous HP and RSA values. The resulting metric describes the observed change in HP (ms) for a one-unit (ln(ms^2^)) change in RSA.

#### Skin conductance response (SCR) frequency

SCRs calculated from the raw EDA signal were used to index sympathetic activation. SCRs are caused by sympathetic activation that promotes the opening of palmar eccrine sweat pores, increasing skin surface conductivity. (35, 61) EDA data were downsampled to 7.812 Hz for analysis and visually inspected. Segments of data with signal noise or disruption were interpolated or excluded from analysis. Phasic data, which were separated from tonic data using smoothing baseline removal, were used to locate SCRs via trapezoidal numeric integration (minimum amplitude criterion = 03 µS) and summed over time to reflect firing rate per minute.

### Data analysis

Statistical analyses were conducted using R 4.1 (42) and RStudio 1.4. (62) Physiological change scores were calculated by subtracting lift and recovery values from baseline. BPQ autonomic reactivity scores were converted to T scores and tertiles based on general population percentiles from Study 1. BMI was calculated from self-reported weight and height (kg/m^2^).

Group differences were evaluated using Chi-Square and Welch’s unequal variance tests. (63) Change scores and the effects of seated leg lifts on HP and RSA were examined with linear mixed models. Standardized mean difference (Cohen’s d) was calculated for pairwise contrasts. (64) Due to skew and floor effects in SCR data, tobit regression and nonparametric repeated measures model were used. (65) Post hoc pairwise comparisons for non-parametric models were conducted using the two-tailed Mann-Whitney U test for between group comparisons and Wilcoxon signed-rank tests for within group comparisons. Rank-biserial correlations were calculated as an effect size for pairwise differences. (66) All statistical tests had alpha set at .05 and were two-sided. Based on their know associations, HP and RSA models were adjusted for age and BMI (75; associations between physiological variables and covariates are in Supplemental Materials).

## Results

The sample was composed of 62 participants. Descriptive statistics are in Table 3. The mean age was 21.85 years (SD = 4.01, range: 18-36), 67.7% were female, and 48.4% had a bachelor’s degree or higher level of education. The mean BMI was 23.84 (SD = 4.53).

**Table 3.**
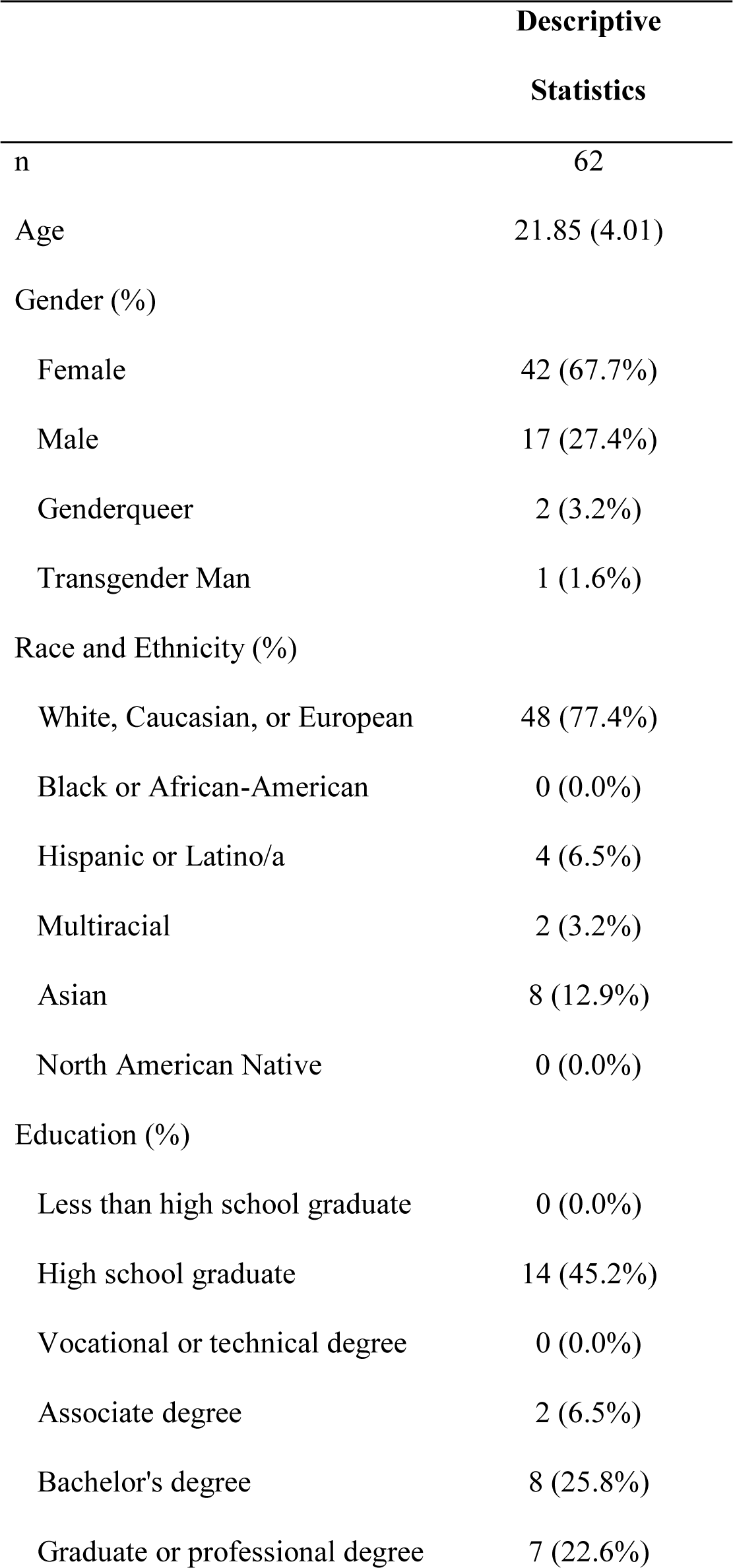
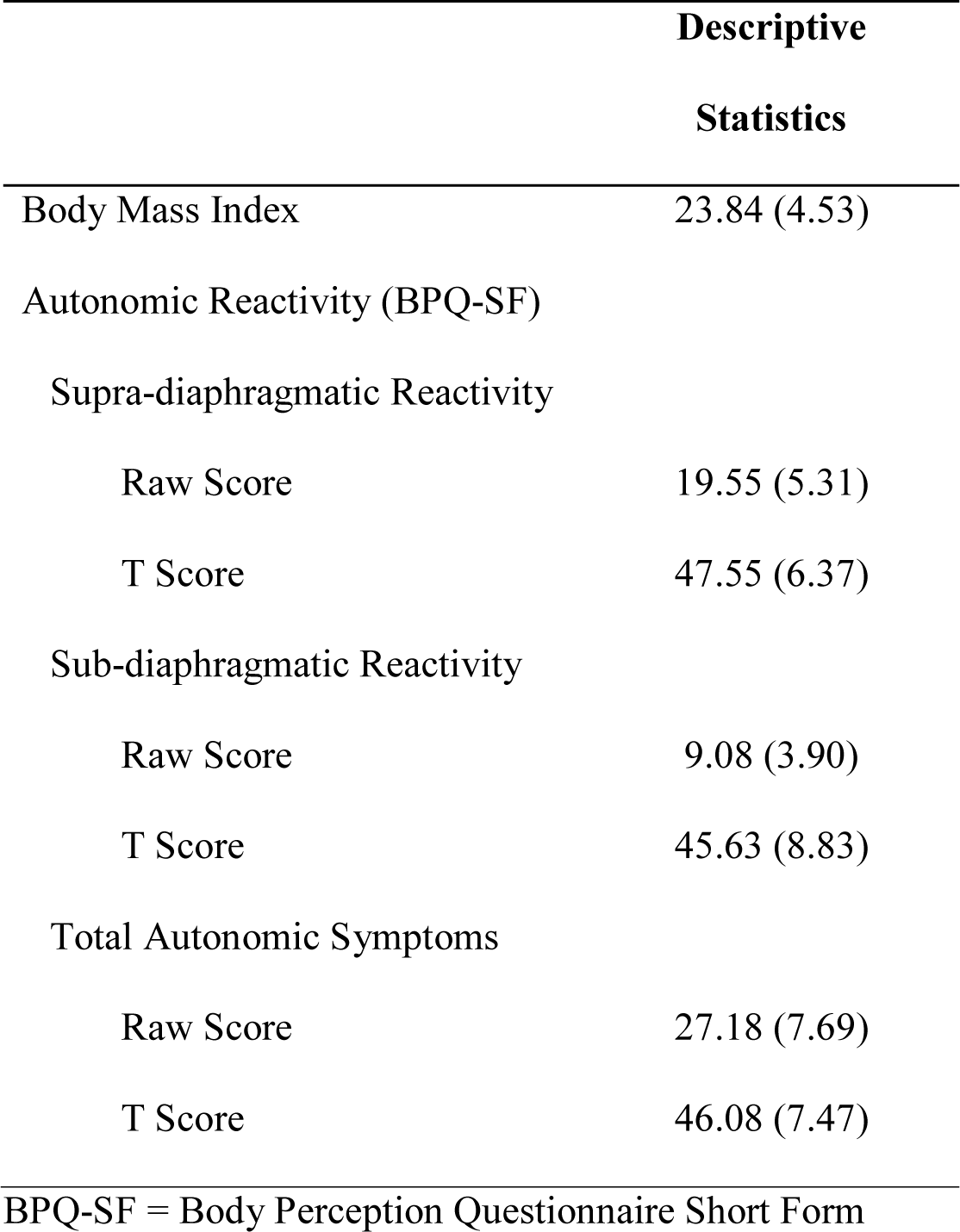
Sample descriptive statistics for Study 2. Continuous variables are summarized with means and standard deviations. Categorical variables are summarized by counts and percentages.

|  | **Descriptive**<br>**Statistics** |
| --- | --- |
| n | 62 |
| Age | 21.85 (4.01) |
| Gender (%) |  |
| Female | 42 (67.7%) |
| Male | 17 (27.4%) |
| Genderqueer | 2 (3.2%) |
| Transgender Man | 1 (1.6%) |
| Race and Ethnicity (%) |  |
| White, Caucasian, or European | 48 (77.4%) |
| Black or African-American | 0 (0.0%) |
| Hispanic or Latino/a | 4 (6.5%) |
| Multiracial | 2 (3.2%) |
| Asian | 8 (12.9%) |
| North American Native | 0 (0.0%) |
| Education (%) |  |
| Less than high school graduate | 0 (0.0%) |
| High school graduate | 14 (45.2%) |
| Vocational or technical degree | 0 (0.0%) |
| Associate degree | 2 (6.5%) |
| Bachelor's degree | 8 (25.8%) |
| Graduate or professional degree | 7 (22.6%) |
| Body Mass Index | 23.84 (4.53) |
| Autonomic Reactivity (BPQ-SF) |  |
| Supra-diaphragmatic Reactivity |  |
| Raw Score | 19.55 (5.31) |
| T Score | 47.55 (6.37) |
| Sub-diaphragmatic Reactivity |  |
| Raw Score | 9.08 (3.90) |
| T Score | 45.63 (8.83) |
| Total Autonomic Symptoms |  |
| Raw Score | 27.18 (7.69) |
| T Score | 46.08 (7.47) |
| BPQ-SF = Body Perception Questionnaire Short Form |  |

Supra- and sub-diaphragmatic scores on the BPQ-SF were positively correlated (*r* = .59) and analysis results were comparable for the two subscales. Thus, results are reported on combined autonomic symptom scores (see Supplemental Materials for supra- and sub-diaphragmatic results reported separately). BPQ-SF autonomic symptom scores from the lab study were lower compared to the general population in Study 1 (low tertile n = 29, 46.8%, mid tertile n = 22, 35.5%, high tertile n = 11, 17.7%). BPQ autonomic symptom scores were not associated with age, gender, body mass index, or meeting overweight and obesity criteria (all p > .05). After data inspection, 9 EDA files were excluded due to signal drop out, poor quality, or equipment errors (final n = 53).

### Associations among physiological variables

Correlations were calculated across the 7 postures (baseline, 3 sitting, 3 recovery). HP and RSA were moderately correlated (Pearson r = .49, p < .001). SCR rate was not significantly correlated with HP (Rho = -.17, p = .226) or RSA (Rho = -.06, p = .68).

### Associations between physiological variables and self-reported autonomic symptoms

BPQ autonomic symptom groups did not differ significantly on HP, RSA, or SCR rate at baseline (all contrasts p > .10). BPQ scores were negatively associated with lower SCR rate during leg lifts (B = -0.24, SE = 0.083, z = -2.94, p = 0.003) and recovery periods (B = - 0.16, SE = 0.05, z = -2.99, p = 0.003), but no other linear associations were significant.

All BPQ autonomic symptom tertile groups reduced mean heart period from baseline to leg lifts (Low = -57.21 [95% CI: -68.04, -46.38], d = 2.41; Mid = -64.53 [95% CI: -76.96, - 52.10], d = 2.72; High = -48.91 [95% CI: -66.49, -31.33], d = 2.06) and there were no significant differences among them during leg lifts (all contrasts p > .30). Only the mid autonomic symptom group maintained shorter mean heart period during recovery conditions compared to baseline (Figure 2 bottom left). This effect was significantly different from the low symptom group, in which the recovery periods were not distinguishable from baseline.

Only the low and mid symptom tertile groups significantly decreased RSA in response to the leg lift task (Low = -.43 [95% CI: -.63, -.23], d = 1.12; Mid = -.53 [95% CI: -.76, -.31], d = 1.37). Both groups returned to baseline levels during recovery periods (Baseline-Recovery Change Low = -.10 [95% CI: -.29, .10], d = 0.25; Mid = -.07 [95% CI: -.29, .16], d = 0.17). The high symptom group did not show significant RSA change from baseline to leg lifts (B = -.15 [95% CI: -.47, .17], d = 0.39) or from baseline to recovery periods (B = -.14 [95% CI: -.18, .46], d = 0.37).

SCR rate increased with leg lifts only in the low and mid groups (Low = 2.47 [95% CI: .82, 4.12], r = -0.60; Mid = 2.50 [95% CI: .65, 4.35], r = -0.71). During recovery periods, the SCR rate significantly declined from leg lifts in the low group (B = 1.70 [95% CI: .69, 2.71], r = 0.58) and was not significantly different from baseline. In contrast, the mid group maintained skin conductance activation above baseline values during recovery periods (Baseline-Recovery B = 2.19 [95% CI: .34, 4.03], r = -0.73). The high symptom group did not significantly differ from baseline during leg lifts or recovery periods. Overall, the high symptom group had a lower SCR rate during leg lifts and recovery compared to the low symptom group (leg lift p = .007; r = .64; recovery p = .002; r = .72), and the mid group was higher than the high group at recovery (p = .002; r = .77). VE was not associated with BPQ autonomic symptom levels (F (2, 53) = .83, p = .442).

## Discussion

This is the first study to conduct factor analysis on full item responses of the English language Body Perception Questionnaire (BPQ-SF), derive normative values for research and clinical use, and assess the convergence of BPQ autonomic reactivity self-reports with sensor-based measures of parasympathetic and sympathetic function. The results demonstrate that 1) the autonomic reactivity subscales have a 1- or 2-factor structure when explored with full item response distributions in a large, general US population of adults; 2) that low levels of self-reported autonomic symptoms are associated with dynamic parasympathetic and sympathetic responses to a metabolic challenge, while high self-reported autonomic symptoms show less dynamically responsive patterns, and 3) that normative values from the general population study can be used as reference values for clinical studies and provide meaningful interpretation of autonomic symptoms.

In study 1, the results of exploratory and confirmatory factor analysis in a large, general population sample converged with prior factor analysis that had been conducted using simplified item response ranges in studies with convenience and student samples. The results revealed two autonomic symptom factors that differentiated supra- and sub-diaphragmatic reactivity. Also consistent with prior studies, the supra- and sub-diaphragmatic subscales were strongly positively correlated (24, 26), suggesting that experiences of autonomic symptoms above and below the diagram are closely associated. In addition, supra- and sub-diaphragmatic reactivity subscale patterns showed similar associations with sensor-based physiological measures in lab data in Study 2. Prior studies have also found patterns of parallel associations between supra- and sub-diaphragmatic symptoms with trauma history, PTSD symptoms, and depression symptoms. (67, 68) This suggests that under some conditions supra- and sub-diaphragmatic reactivity may be scored as a single subscale, though distinctions between subscales should continue to be explored. It also underscores that processes that regulate organs above and below the diagram are functionally integrated and likely impacted in tandem.

Consistent with previous studies, we found that the urge to vomit is associated with both supra- and sub-diaphragmatic reactivity factors (24, 26), likely reflecting the neuro-anatomical circuits that induce vomiting, which involve structures both above and below the diaphragm. (45, 69) Vomiting requires a coordinated outflow from both the dorsal motor nucleus of the vagus (which mainly innervates structures below the diaphragm) and the nucleus ambiguus (regulating structures above the diaphragm) which synchronize swallowing, respiration, cardiovascular activity, and gastrointestinal function. (69) Disruption at any of these pathways may result in sensitization towards an urge to vomit.

In study two, participants in all autonomic symptom level groups responded metabolically to leg lifts with shortened average heart period. However, parasympathetic and sympathetic metrics showed different patterns of dynamic autonomic regulation associated with self-reported autonomic symptoms. Those with low autonomic symptoms had the most flexible autonomic regulation, withdrawing PNS and activating SNS during leg lifts, then increasing PNS while reducing SNS rapidly between challenges. This pattern reflects maximum flexibility in responding to momentary metabolic needs, a hallmark of efficient allostasis. (6, 38) This dynamic flexibility may protect individuals from extended or uncontrolled activation that lasts beyond immediate metabolic needs which could lead to experiencing autonomic symptoms.

Similarly, moderate self-reported autonomic symptom levels were also associated with efficient withdrawal and re-engagement of the parasympathetic vagal brake. However, while this group exhibited sympathetic activation during leg lifts compared to baseline, this sympathetic activation was maintained even during rest periods. This was also reflected in average heart period change, where HP was shortened even during rest periods. Lack of dynamic flexibility in reducing sympathetic responses may cause some disruption in organ function over time, resulting in more experiences of autonomic symptoms.

The highest levels of self-reported autonomic symptoms were associated with no consistent parasympathetic or sympathetic change. Individuals at these levels may be least organized in their autonomic response, not predictably engaging or disengaging the ventral vagal parasympathetic brake or the SNS, placing them most at risk of long-term disruptions in the function of organs and tissues innervated by the autonomic nervous system. However, this group nonetheless mounted a change in overall heart period to leg lifts and a return toward baseline during rest periods, a pattern that requires some neural regulation of the heart. The polyvagal theory posits that metabolic regulation via the unmyelinated cardioinhibitory vagal pathway to the heart via the dorsal vagal motor nucleus of the vagus (DMNX) would be associated with the highest levels of autonomic symptoms due to its being the evolutionarily oldest pathway and its intimate integration with the gut. (25) Electrophysiological studies have documented that direct stimulation of the DMNX will produce heart rate slowing (70–72) and that vagal activation influences sympathetic outflow (e.g., (73)). However, there is no known non-invasive marker of the cardioinhibitory influence of the DMNX and the data in this study cannot confirm whether it is involved in cardiac regulation during leg lifts in the high symptom group.

VE was not associated with autonomic symptoms in this study. Prior studies have quantified VE during posture shifts that engage the baroreflex maintaining blood pressure to the brain (37, 60) or strenuous exercise on a stationary bicycle that requires strenuous metabolic muscle activation. (74) Compared to these studies, leg lifts may be a relatively weak ANS challenge, not able to provide enough metabolic engagement to permit robust VE measurement. Future studies that use paradigms that provide stronger challenge are needed.

Viewed in sum, these studies support the use of self-reported autonomic symptoms via the BPQ as an index of autonomic disruption of typical organ and tissue function. However, there are important distinctions between sensor-based measures, which are typically used to index autonomic activity, and self-reports. First, self-reports and sensor-based measures differ on their ultimate goals. Whereas sensors typically focus on the dynamic function of a single organ or tissue, self-reports pool information over many innervation targets. Also, self-reported symptoms are likely most noticeable when their function is disrupted and are likely less sensitive to slight fluctuations in dynamic neural regulation. In addition, self-reports concatenate experience over long periods of time to provide enough opportunity to observe potential disruptions. Finally, compared to sensor-based measures, questionnaires have measurement limitations that include reliance on memory, reporting biases, attention, and other cognitive factors. However, the benefits of self-report measures in providing an overall summary of autonomic disruption at a low burden for participants makes them a potentially useful part of an assessment battery.

Though the BPQ may index disruptions in regulation of the autonomic nervous system, the established factor structure is not specific to a particular autonomic branch. While some organs and tissues are innervated by a single pathway, such as the sympathetic regulation of sweat glands in the palm, most autonomic innervation targets are regulated by multiple branches. (5, 11) These multiple innervation paths, along with the co-regulation of the various branches of the ANS by higher level structures such as the hypothalamus, (5) make distinctions between specific autonomic pathways challenging for self-report measurement. BPQ-SF autonomic symptoms consistently cluster together across samples without specificity to one branch or another. The findings reported here, however, do support the use of the BPQ-SF autonomic symptom items as a measure of allostatic flexibility or autonomic resilience. High symptoms are associated with weaker flexibility to mount responses to challenge and recover after metabolic resources are not needed.

Clinical and research use of self-report symptoms can be supported by the derivation of normative values and generation of cut scores. This paper describes the first derivation of normative percentile and T scores from a large general US sample. While study 2 used a prior division into tertiles, showing non-linear distinctions between low, moderate, and high symptom groupings, larger samples are needed to optimize cut scores to capture autonomic disruption with optimized sensitivity and specificity.

### Limitations

Study 2 was based on a community convenience sample. We found that the sample had restricted age and BMI distribution, compared to a general population. Both factors have a known association with patterns of autonomic function. (75) This sample also had a larger proportion of low autonomic symptom scores and lower proportion of those at the highest levels compared to the online sample that was more representative of the range of BMI and age in a general population. Future studies need to examine the association with autonomic symptoms with sensor-based measures in more broad population-based samples.

## Conclusion

These studies support the use of the BPQ-SF autonomic reactivity subscales and their validity to index autonomic disruption of organ functions. Assessing experiences of autonomic symptoms can be utilized in clinical monitoring of patient symptoms and in basic research into mechanisms that affect mental wellbeing and physical health.

## Data Availability

All data produced in the present study are available upon reasonable request to the authors

## Acknowledgements

We wish to thank Juliet Dahlgren for assistance with physiological data processing and R. Mike Winters on an early version of physiological data analysis.

## ACRONYMS

ANS: autonomic nervous system
BMI: body mass index
BPQ: Body Perception Questionnaire
BPQ-SF: Body Perception Questionnaire
Short Form CFA: confirmatory factor analysis
CFI: Comparative Fit Index
DMNX: dorsal motor nucleus of the vagus
ECG: electrocardiogram
EDA: electrodermal activity
EFA: exploratory factor analysis
FIR: finite impulse response
HF-HRV: high frequency heart rate variability
IBI: inter-beat intervals
HP: heart period
NA: nucleus ambiguus
PNS: parasympathetic nervous system
PTSD: post-traumatic stress disorder
RMSEA: root mean squared error of approximation
RSA: respiratory sinus arrythmia
SCR: skin conductance reactivity
SLR: seated straight-leg raise
SNS: sympathetic nervous system
TLI: Tucker-Lewis Index
VE: vagal efficiency
VVC: ventral vagal complex

