## Supplemental Material for "Measuring Autonomic Symptoms with the Body Perception Questionnaire Short Form (BPQ-SF): Factor Analysis, Derivation of U.S. Adult Normative Values, and Association with Sensor-Based Physiological Measures"

Supplemental Materials

Study 1

*Sample recruitment and data preparation*. Data were collected in August and September 2018. The recruitment database is drawn and maintained through various sources, including website intercept recruitment, member referrals, targeted email lists, gaming sites, and customer loyalty web portals. Consumer panel members’ names, addresses, and dates of birth are validated via a third party prior to their joining a panel but were not used or accessed by the researchers in this study. Individuals in the database were sent an email invitation or prompted on the survey platform. Participant payment was conducted by the specific source after the survey was completed. The payment amount and method (e.g., SkyMiles, cash, gift cards) were determined by that participant's individual plan with compensation typically ranging between $2.50-4.00.

Data quality was assessed by automated checks and manual inspection. Responses with large sections of identical responses for any one survey section were checked for internal consistency and compared to item response patterns in prior studies. Responses that did not meet criteria for high quality or had a response time completion time faster than 1/3 of the median completion time were excluded. Exploratory factor analysis (EFA) was conducted on body awareness and autonomic reactivity domains separately (Factor Analysis results on body awareness can be found in Supplemental Materials). Missing values comprised less than 8% of any one item and less than 5% of all data. Inspection of missing values did not indicate any systematic patterns of missingness.

*BPQ exploratory factor analysis*. Parallel analysis for the awareness subscale showed evidence for a possible 1-3 factors (Figure S1). The plot was dominated by one prominent eigenvalue, and two additional eigenvalues that weakly diverged from the simulated and resampled data. TLI and RMSEA approached good fit in the one-factor solution but did not reach our criteria for good fit until the third factor was added to the model. Upon inspection of the loadings, the 3-factor solution evidenced overfitting the data by violating simple structure, with several factors having substantial loadings on the same items and, in the 3 to 5- factor solutions, some items lacking any substantial loadings (Table S1 and Figures S2 and S3). The 5-factor solution did not converge. The 2-factor solution was not interpretable and had a high correlation (r = .85) between factors. Given the prominence of the single eigenvalue in the parallel analysis and the simple structure generated by the one-factor solution, the one-factor model was retained for the CFA. Geomin rotated standardized loadings using this solution ranged from .62 to .86.

The autonomic reactivity EFA scree plot showed a substantial drop after the first eigenvalue, and a more moderate drop after the second eigenvalue, before leveling out and converging with the simulated and resampled data, supporting a one or two factor solution (Figure S1). The RMSEA approached good fit for the one- and two-factor solutions, but did not meet our criteria for good fit until the four-factor solution. However, solutions with more than 2 factors violated simple structure, resulting in factors that had substantial loadings on multiple items, items that substantially loaded on only one factor, and items that did not load strongly on any factor. These patterns suggested overfitting the data. Thus, the three- and four-factor solution were discarded as improbable. As found in prior factor analyses, the two-factor solution had one item that loaded substantially onto both factors (“I feel like vomiting”) and the rest of the items demonstrated a simple structure by only loading strongly to one of the factors. Geomin rotated standardized loadings for 2-factor solution were as follows: factor 1 [range: 0.4 - 0.94] factor 2 [range: 0.47 – 0.93].

Exploratory factor analysis. Parallel analysis and fit indices suggested 1-3 factor solutions for body awareness. Examination of factor loadings showed that solutions with greater than 1 body awareness factor violated simple structure, with several factors having strong loadings on the same items and some lacking any substantial loadings. Thus, the 1-factor body awareness solution was retained for confirmatory factor analysis

Confirmatory factor analysis. The EFA results were tested using CFA on the other half of the split sample. This structure fit well in the Awareness domain (*RMSEA* = .070 [90% CI: .067, .074], *CFI* = .99, *TLI* = .99). CFA Loadings were similar to those found in the EFA (Table 2 and 3).

Figure S1. Parallel analysis plots

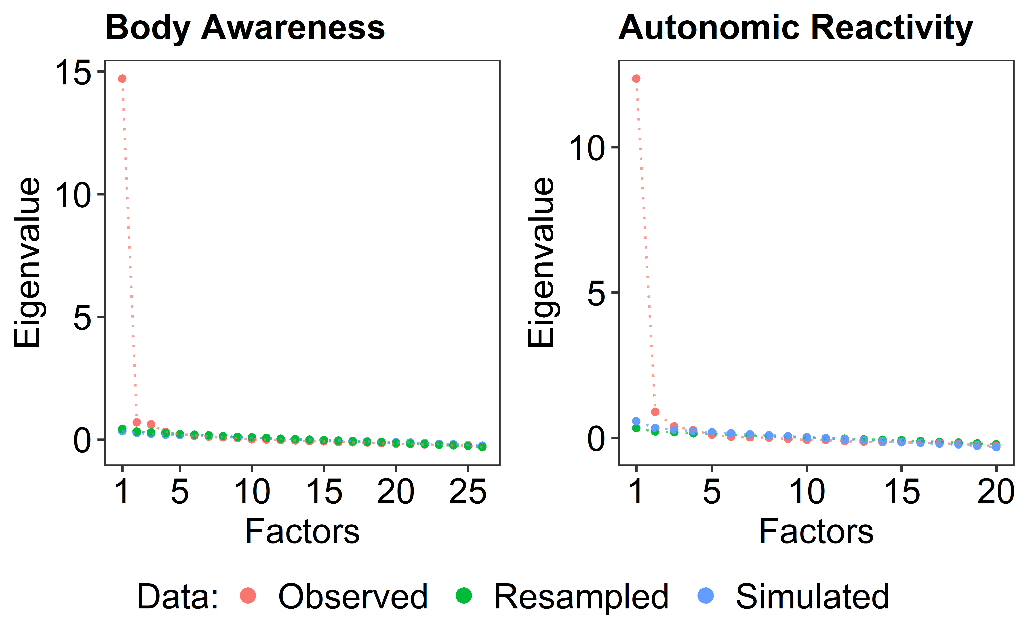

Table S1. Exploratory factor analysis fit indices

| **Domain** | **Factors** | **DF** | **Chi Square** | **CFI** | **TLI** | **RMSEA** | | |
| --- | --- | --- | --- | --- | --- | --- | --- | --- |
|  |  |  |  |  |  | **Estimate** | **Lower** | **Upper** |
| Body Awareness | 1 | 299 | 1548.422 | 0.852 | 0.839 | 0.104 | 0.101 | 0.107 |
|  | 2 | 274 | 1002.352 | 0.882 | 0.86 | 0.097 | 0.094 | 0.1 |
|  | 3 | 250 | 538.512 | 0.923 | 0.9 | 0.082 | 0.078 | 0.085 |
|  | 4 | 227 | 388.268 | 0.939 | 0.913 | 0.076 | 0.073 | 0.08 |
|  | 5 | 205 | 291.595 | 0.953 | 0.925 | 0.071 | 0.067 | 0.074 |
| Autonomic Reactivity | 1 | 170 | 1401.104 | 0.832 | 0.812 | 0.138 | 0.135 | 0.143 |
|  | 2 | 151 | 460.091 | 0.907 | 0.882 | 0.109 | 0.105 | 0.114 |
|  | 3 | 133 | 253.266 | 0.936 | 0.909 | 0.096 | 0.092 | 0.101 |
|  | 4 | 116 | 135.779 | 0.96 | 0.935 | 0.081 | 0.076 | 0.086 |
|  | 5 | 100 | 95.205 | 0.967 | 0.937 | 0.08 | 0.074 | 0.085 |

DF = Degrees of freedom

Table S2. Body Awareness subscale loadings from exploratory factor analysis (EFA) and confirmatory factor analysis (CFA)

| **Item Number** | **Item** | **EFA** | **CFA** |
| --- | --- | --- | --- |
| 1 | Swallowing frequently | 0.6 | 0.66 |
| 2 | An urge to cough to clear my throat | 0.64 | 0.66 |
| 3 | My mouth being dry | 0.66 | 0.67 |
| 4 | How fast I am breathing | 0.74 | 0.76 |
| 5 | Watering or tearing of my eyes | 0.73 | 0.74 |
| 6 | Noises associated with my digestion | 0.74 | 0.75 |
| 7 | A swelling of my body or parts of my body | 0.76 | 0.78 |
| 8 | An urge to defecate | 0.64 | 0.64 |
| 9 | Muscle tension in my arms and legs | 0.76 | 0.8 |
| 10 | A bloated feeling because of water retention | 0.77 | 0.83 |
| 11 | Muscle tension in my face | 0.83 | 0.82 |
| 12 | Goose bumps | 0.76 | 0.79 |
| 13 | Stomach and gut pains | 0.83 | 0.86 |
| 14 | Stomach distension or bloatedness | 0.81 | 0.88 |
| 15 | Palms sweating | 0.77 | 0.81 |
| 16 | Sweat on my forehead | 0.74 | 0.78 |
| 17 | Tremor in my lips | 0.86 | 0.88 |
| 18 | Sweat in my armpits | 0.69 | 0.75 |
| 19 | The temperature of my face (especially my ears) | 0.8 | 0.81 |
| 20 | Grinding my teeth | 0.69 | 0.75 |
| 21 | General jitteriness | 0.79 | 0.84 |
| 22 | The hair on the back of my neck ""standing up"" | 0.77 | 0.78 |
| 23 | Difficulty in focusing | 0.73 | 0.75 |
| 24 | An urge to swallow | 0.82 | 0.8 |
| 25 | How hard my heart is beating | 0.81 | 0.81 |
| 26 | Feeling constipated | 0.75 | 0.76 |

Figure S2. Exploratory factor analysis loadings for BPQ-SF body awareness

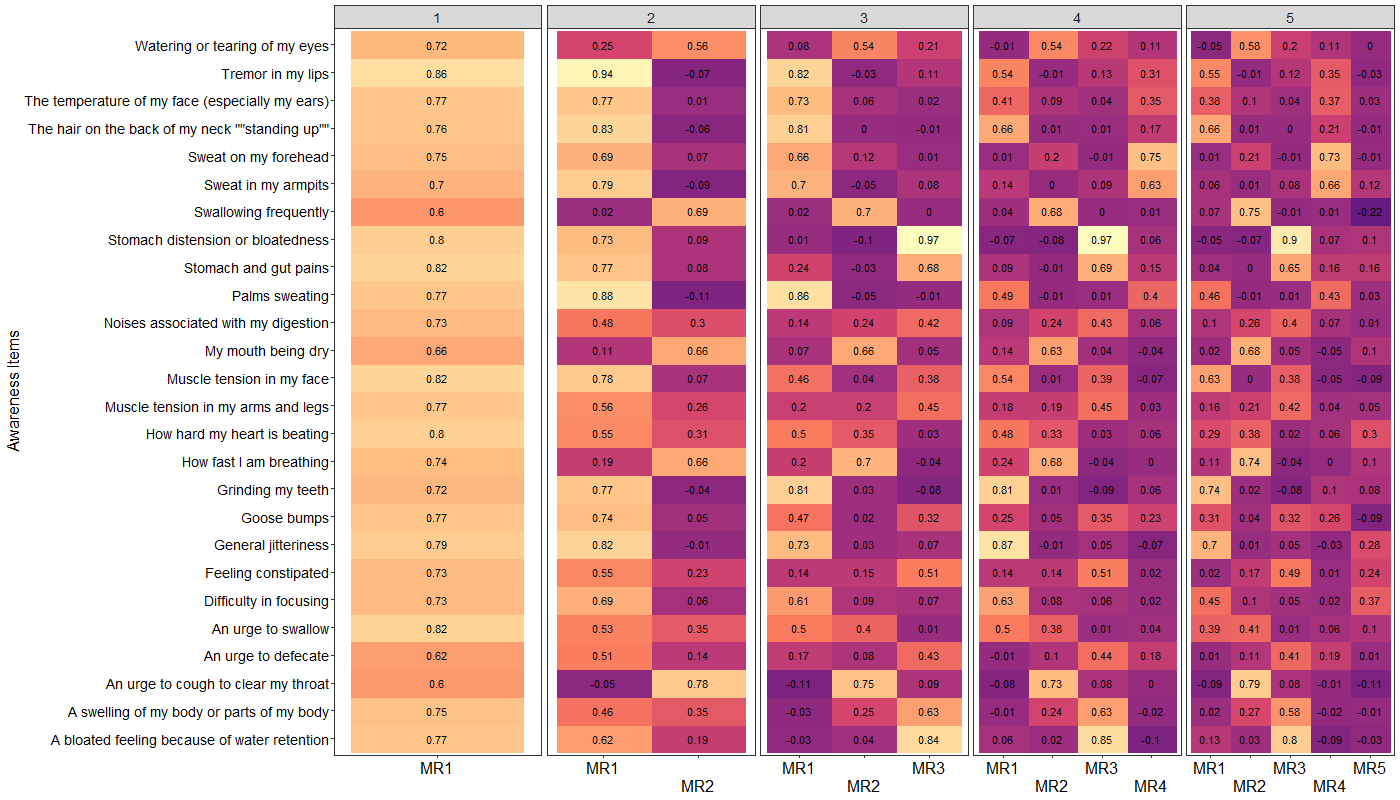

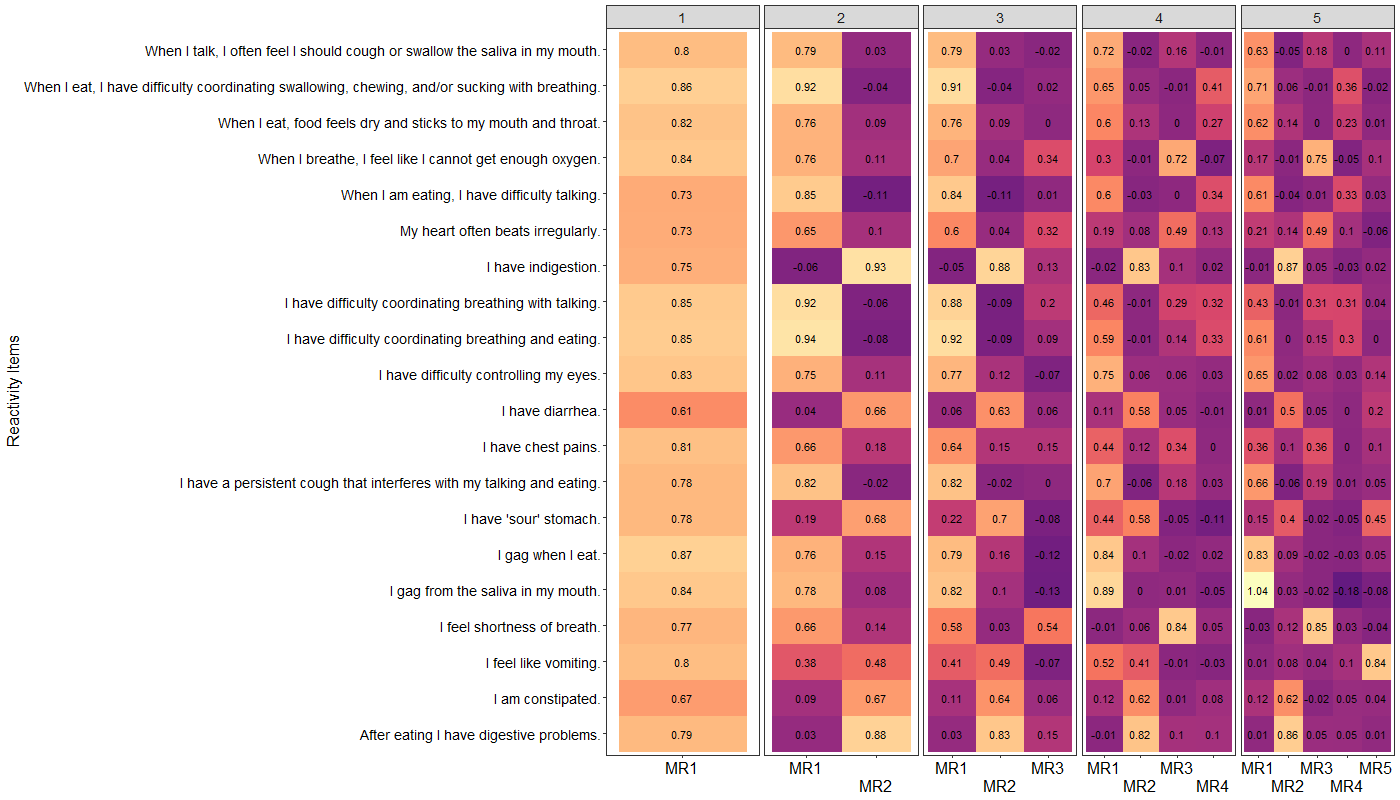
Figure S3. Exploratory factor analysis loadings for BPQ-SF autonomic reactivity items

Tables S3. BPQ-SF EFA ANS reactivity subscale factor correlations

2 Factor Correlations

|  | **MR1** | **MR2** |
| --- | --- | --- |
| **MR1** | 1 | 0.79 |
| **MR2** | 0.79 | 1 |

3 Factor Correlations

|  | **MR1** | **MR2** | **MR3** |
| --- | --- | --- | --- |
| **MR1** | 1 | 0.787 | 0.1 |
| **MR2** | 0.787 | 1 | 0.116 |
| **MR3** | 0.1 | 0.116 | 1 |

4 Factor Correlations

|  | **MR1** | **MR4** | **MR2** | **MR3** |
| --- | --- | --- | --- | --- |
| **MR1** | 1 | 0.89 | 0.669 | 0.73 |
| **MR2** | 0.89 | 1 | 0.638 | 0.779 |
| **MR3** | 0.669 | 0.638 | 1 | 0.627 |
| **MR4** | 0.73 | 0.779 | 0.627 | 1 |

5 Factor Correlations

|  | **MR1** | **MR4** | **MR2** | **MR3** | **MR5** |
| --- | --- | --- | --- | --- | --- |
| **MR1** | 1 | 0.906 | 0.682 | 0.726 | 0.044 |
| **MR2** | 0.906 | 1 | 0.654 | 0.771 | 0.055 |
| **MR3** | 0.682 | 0.654 | 1 | 0.62 | 0.287 |
| **MR4** | 0.726 | 0.771 | 0.62 | 1 | 0.187 |
| **MR5** | 0.044 | 0.055 | 0.287 | 0.187 | 1 |

Figure S4. BPQ-SF raw item distributions

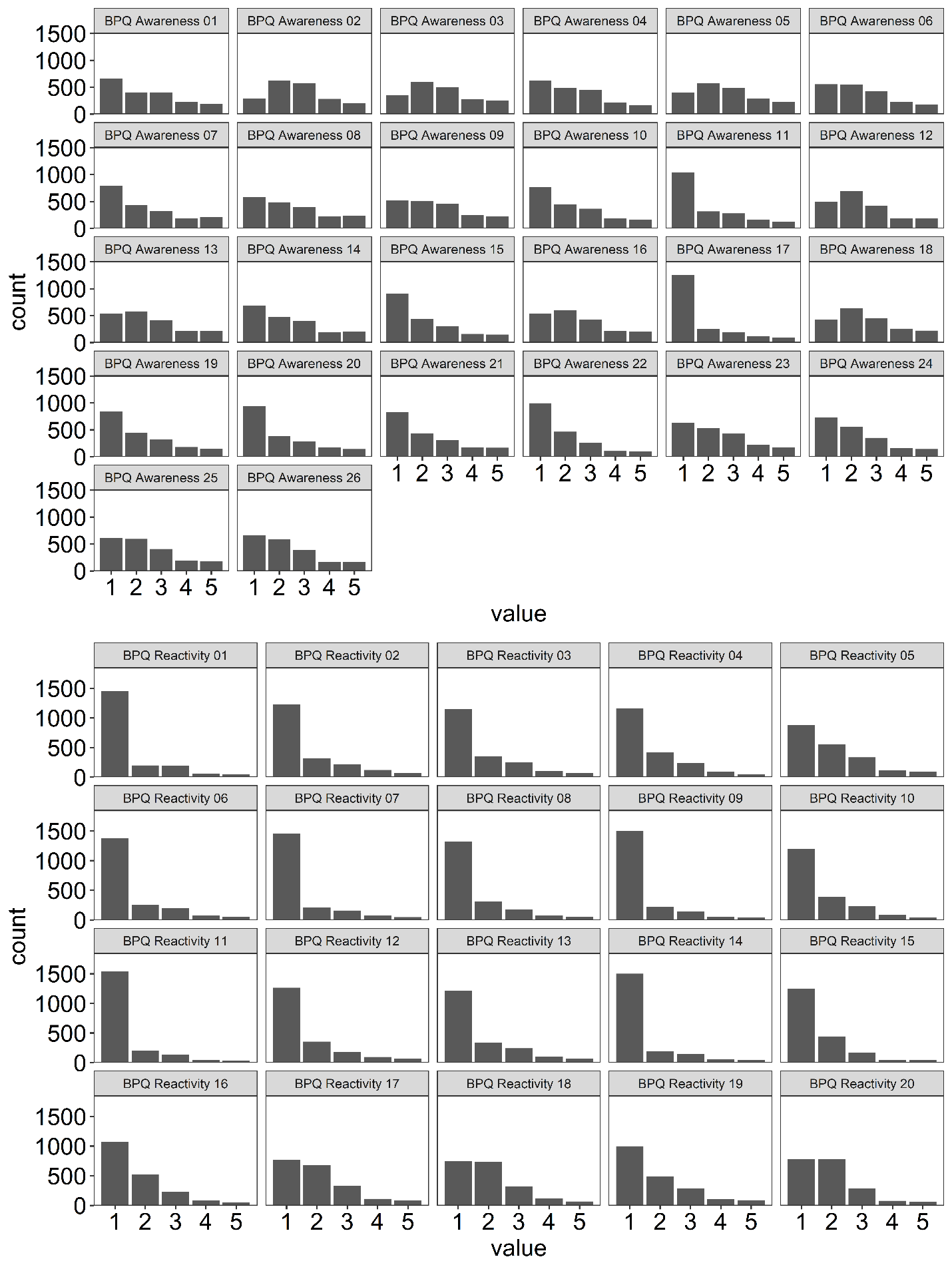

*Calculation of percentiles and T scores.* Nearly all possible subscale scores were observed in the data. Three supra-diaphragmatic reactivity subscale scores that were possible based on combinations of items but not observed were interpolated. Percentile rank and T scores are available for raw scores computed by the sum of full item responses. These transformed scores are based on a combined sample of American participants recruited online. Participant age ranged from 18 to 90 years (Mean = 46.6, SD = 17.2). 49.5% of the sample were men, 49.2% were women, and 0.9% were transgender or genderqueer. 80 individuals did not provide age, while 9 did not provide a gender. Age- and gender-specific norms are not yet available. To transform raw values to percentile ranks and T-scores, use the tables on the next pages. To mitigate the influence of missing values, missing values were set to the participant median of non-missing responses and participants with more than 5 missing values were excluded (191; 9% of the sample).

***Percentile rank scores*** reflect the percentage of values that are equal or lower to the individual’s score. For example, a score in the 5^th^ percentile rank means that the individual’s score is greater or equal to 5% of the scores in a normative sample. A 95^th^ percentile rank means that the individual’s score is greater or equal to 95% of the scores in a normative sample.

***T scores*** reflect a standardized value according to a normal distribution based on a mean of 50 and a standard deviation of 10. This transformation is recommended for parametric statistical models.

To account for possible raw scores that no participant received for a given subscale, the scores that were obtained were used to generate a monotonic Hermite spline function, from which approximate percentile ranks and T scores were generated. Such scores are marked with an asterisk in the tables on the following pages. R code for this process and for standard percentile rank and T score transformations is available by request.

Table S4. Body awareness raw scores, percentile ranks, and T scores

**Body Awareness**

| *Raw Score* | *Percentile Rank* | *T-Score* | *Raw Score* | *Percentile Rank* | *T-Score* |
| --- | --- | --- | --- | --- | --- |
| *26* | 1.3% | 27.7 | ***63*** | 62.2% | 53.1 |
| *27* | 2.9% | 31.0 | ***64*** | 63.7% | 53.5 |
| *28* | 3.6% | 32.0 | ***65*** | 65.2% | 53.9 |
| *29* | 4.7% | 33.2 | ***66*** | 66.7% | 54.3 |
| *30* | 5.7% | 34.2 | ***67*** | 68.0% | 54.7 |
| *31* | 6.9% | 35.2 | ***68*** | 69.1% | 55.0 |
| *32* | 8.3% | 36.2 | ***69*** | 70.3% | 55.3 |
| *33* | 9.7% | 37.0 | ***70*** | 71.6% | 55.7 |
| *34* | 11.0% | 37.8 | ***71*** | 73.0% | 56.1 |
| *35* | 12.5% | 38.5 | ***72*** | 74.1% | 56.5 |
| *36* | 14.3% | 39.3 | ***73*** | 75.0% | 56.7 |
| *37* | 16.3% | 40.2 | ***74*** | 75.9% | 57.0 |
| *38* | 18.3% | 41.0 | ***75*** | 77.0% | 57.4 |
| *39* | 20.2% | 41.7 | ***76*** | 77.9% | 57.7 |
| *40* | 21.9% | 42.3 | ***77*** | 78.6% | 57.9 |
| *41* | 23.7% | 42.9 | ***78*** | 79.6% | 58.3 |
| *42* | 25.7% | 43.5 | ***79*** | 80.6% | 58.6 |
| *43* | 27.9% | 44.1 | ***80*** | 81.2% | 58.9 |
| *44* | 29.8% | 44.7 | ***81*** | 81.7% | 59.1 |
| *45* | 31.5% | 45.2 | ***82*** | 82.3% | 59.3 |
| *46* | 33.2% | 45.7 | ***83*** | 83.0% | 59.6 |
| *47* | 35.1% | 46.2 | ***84*** | 83.9% | 59.9 |
| *48* | 37.0% | 46.7 | ***85*** | 84.4% | 60.1 |
| *49* | 38.7% | 47.1 | ***86*** | 84.9% | 60.3 |
| *50* | 40.5% | 47.6 | ***87*** | 85.5% | 60.6 |
| *51* | 42.5% | 48.1 | ***88*** | 86.2% | 60.9 |
| *52* | 44.4% | 48.6 | ***89*** | 86.8% | 61.2 |
| *53* | 46.1% | 49.0 | ***90*** | 87.3% | 61.4 |
| *54* | 48.0% | 49.5 | ***91*** | 88.0% | 61.7 |
| *55* | 49.7% | 49.9 | ***92*** | 88.6% | 62.0 |
| *56* | 51.2% | 50.3 | ***93*** | 89.0% | 62.2 |
| *57* | 53.0% | 50.7 | ***94*** | 89.3% | 62.4 |
| *58* | 54.6% | 51.1 | ***95*** | 89.6% | 62.6 |
| *59* | 56.1% | 51.5 | ***96*** | 90.0% | 62.8 |
| *60* | 57.6% | 51.9 | ***97*** | 90.5% | 63.1 |
| *61* | 59.2% | 52.3 | ***98*** | 91.1% | 63.4 |
| *62* | 60.8% | 52.7 | ***99*** | 91.5% | 63.8 |

**Body Awareness (Cont.)**

| *Raw Score* | *Percentile Rank* | *T-Score* | *Raw Score* | *Percentile Rank* | *T-Score* |
| --- | --- | --- | --- | --- | --- |
| *100* | 91.9% | 64.0 | ***116*** | 97.5% | 69.6 |
| *101* | 92.1% | 64.1 | ***117*** | 97.7% | 69.9 |
| *102* | 92.6% | 64.4 | ***118*** | 97.9% | 70.3 |
| *103* | 93.0% | 64.8 | ***119*** | 98.1% | 70.7 |
| *104* | 93.5% | 65.1 | ***120*** | 98.2% | 71.0 |
| *105* | 93.9% | 65.5 | ***121*** | 98.3% | 71.3 |
| *106* | 94.3% | 65.8 | ***122**** | 98.4% | 71.4 |
| *107* | 94.6% | 66.1 | ***123*** | 98.5% | 71.6 |
| *108* | 95.0% | 66.5 | ***124*** | 98.5% | 71.8 |
| *109* | 95.5% | 67.0 | ***125*** | 98.8% | 72.5 |
| *110* | 95.9% | 67.3 | ***126*** | 99.1% | 73.7 |
| *111* | 96.1% | 67.6 | ***127*** | 99.3% | 74.6 |
| *112* | 96.3% | 67.9 | ***128*** | 99.4% | 75.3 |
| *113* | 96.6% | 68.3 | ***129*** | 99.5% | 76.1 |
| *114* | 96.9% | 68.7 | ***130*** | 99.8% | 79.0 |
| *115* | 97.2% | 69.2 |  |  |  |

Table S5. Body awareness very short form raw scores, percentile ranks, and T scores

**Body Awareness Very Short Form (VSF)**

| *Raw Score* | *Percentile Rank* | *T-Score* | *Raw Score* | *Percentile Rank* | *T-Score* |
| --- | --- | --- | --- | --- | --- |
| *12* | 1.6% | 28.7 | ***37*** | 78.5% | 57.9 |
| *13* | 3.9% | 32.4 | ***38*** | 80.2% | 58.5 |
| *14* | 5.5% | 34.0 | ***39*** | 81.9% | 59.1 |
| *15* | 7.5% | 35.6 | ***40*** | 83.5% | 59.7 |
| *16* | 9.7% | 37.0 | ***41*** | 84.9% | 60.3 |
| *17* | 12.4% | 38.5 | ***42*** | 86.3% | 60.9 |
| *18* | 15.7% | 39.9 | ***43*** | 87.4% | 61.5 |
| *19* | 19.6% | 41.5 | ***44*** | 88.3% | 61.9 |
| *20* | 23.6% | 42.8 | ***45*** | 89.3% | 62.4 |
| *21* | 27.1% | 43.9 | ***46*** | 90.3% | 63.0 |
| *22* | 30.7% | 45.0 | ***47*** | 91.1% | 63.5 |
| *23* | 34.9% | 46.1 | ***48*** | 92.5% | 64.4 |
| *24* | 38.9% | 47.2 | ***49*** | 93.9% | 65.4 |
| *25* | 42.2% | 48.0 | ***50*** | 94.7% | 66.2 |
| *26* | 45.6% | 48.9 | ***51*** | 95.4% | 66.9 |
| *27* | 48.9% | 49.7 | ***52*** | 96.0% | 67.6 |
| *28* | 52.3% | 50.6 | ***53*** | 96.6% | 68.3 |
| *29* | 55.7% | 51.4 | ***54*** | 97.2% | 69.2 |
| *30* | 58.8% | 52.2 | ***55*** | 97.6% | 69.8 |
| *31* | 61.6% | 53.0 | ***56*** | 97.9% | 70.3 |
| *32* | 64.5% | 53.7 | **57** | 98.3% | 71.2 |
| *33* | 67.7% | 54.6 | ***58*** | 98.8% | 72.4 |
| *34* | 70.8% | 55.5 | ***59*** | 99.1% | 73.6 |
| *35* | 73.6% | 56.3 | ***60*** | 99.6% | 76.7 |
| *36* | 76.3% | 57.2 |  |  |  |

Table S6. Supra-diaphragmatic reactivity raw scores, percentile ranks, and T scores

**Supradiaphragmatic Reactivity**

| *Raw Score* | *Percentile Rank* | *T-Score* | *Raw Score* | *Percentile Rank* | *T-Score* |
| --- | --- | --- | --- | --- | --- |
| *15* | 9.3% | 36.8 | **46** | 93.8% | 65.4 |
| *16* | 23.6% | 42.8 | **47** | 94.1% | 65.7 |
| *17* | 32.8% | 45.5 | ***48*** | 94.5% | 66.0 |
| *18* | 40.1% | 47.5 | **49** | 94.9% | 66.4 |
| *19* | 46.3% | 49.1 | **50** | 95.2% | 66.7 |
| *20* | 51.6% | 50.4 | ***51*** | 95.4% | 66.9 |
| *21* | 56.3% | 51.6 | ***52*** | 95.7% | 67.2 |
| *22* | 60.7% | 52.7 | ***53*** | 96.0% | 67.5 |
| *23* | 64.7% | 53.8 | **54** | 96.2% | 67.8 |
| *24* | 68.0% | 54.7 | **55** | 96.6% | 68.2 |
| *25* | 70.6% | 55.4 | **56** | 97.0% | 68.8 |
| *26* | 72.9% | 56.1 | **57** | 97.3% | 69.3 |
| *27* | 74.4% | 56.6 | **58** | 97.5% | 69.6 |
| *28* | 76.0% | 57.1 | **59** | 97.7% | 69.9 |
| *29* | 78.0% | 57.7 | ***60*** | 98.0% | 70.5 |
| *30* | 79.6% | 58.3 | ***61*** | 98.3% | 71.1 |
| *31* | 81.0% | 58.8 | ***62**** | 98.4% | 71.5 |
| *32* | 82.6% | 59.4 | **63** | 98.6% | 71.9 |
| *33* | 83.9% | 59.9 | **64** | 98.8% | 72.7 |
| *34* | 85.0% | 60.4 | **65** | 99.0% | 73.2 |
| *35* | 85.9% | 60.7 | **66** | 99.0% | 73.4 |
| *36* | 86.7% | 61.1 | **67** | 99.1% | 73.7 |
| *37* | 87.4% | 61.5 | **68** | 99.2% | 74.3 |
| *38* | 88.3% | 61.9 | **69** | 99.4% | 75.3 |
| *39* | 89.2% | 62.4 | **70** | 99.6% | 76.3 |
| *40* | 89.8% | 62.7 | **71** | 99.6% | 76.7 |
| *41* | 90.4% | 63.1 | **72*** | 99.6% | 76.9 |
| *42* | 91.2% | 63.5 | **73** | 99.7% | 77.2 |
| *43* | 91.8% | 63.9 | **74*** | 99.8% | 78.2 |
| *44* | 92.4% | 64.3 | **75** | 99.9% | 80.0 |
| *45* | 93.1% | 64.8 |  |  |  |

Table S7. Sub-diaphragmatic reactivity raw scores, percentile ranks, and T scores

**Subdiaphragmatic Reactivity**

| *Raw Score* | *Percentile Rank* | *T-Score* |
| --- | --- | --- |
| *6* | 8.9% | 36.5 |
| *7* | 22.0% | 42.3 |
| *8* | 31.3% | 45.1 |
| *9* | 40.4% | 47.6 |
| *10* | 49.7% | 49.9 |
| *11* | 59.2% | 52.3 |
| *12* | 68.0% | 54.7 |
| *13* | 74.6% | 56.6 |
| *14* | 78.9% | 58.0 |
| *15* | 82.5% | 59.3 |
| *16* | 85.8% | 60.7 |
| *17* | 88.6% | 62.0 |
| *18* | 90.5% | 63.1 |
| *19* | 92.2% | 64.2 |
| *20* | 93.4% | 65.1 |
| *21* | 94.6% | 66.1 |
| *22* | 95.6% | 67.1 |
| *23* | 96.4% | 68.0 |
| *24* | 97.2% | 69.2 |
| *25* | 97.8% | 70.2 |
| *26* | 98.5% | 71.6 |
| *27* | 99.0% | 73.4 |
| *28* | 99.4% | 75.0 |

Table S8. Combined reactivity (supradiaphagmatic and subdiaphragmatic) raw scores, percentile ranks, and T scores

**Combined Reactivity**

| *Raw Score* | *Percentile Rank* | *T-Score* | *Raw Score* | *Percentile Rank* | *T-Score* |
| --- | --- | --- | --- | --- | --- |
| *21* | 5.2% | 33.8 | ***58*** | 90.7% | 63.2 |
| *22* | 12.9% | 38.7 | ***59*** | 91.2% | 63.5 |
| *23* | 18.1% | 40.9 | ***60*** | 91.7% | 63.8 |
| *24* | 23.5% | 42.8 | ***61*** | 92.1% | 64.1 |
| *25* | 28.9% | 44.4 | ***62*** | 92.6% | 64.5 |
| *26* | 33.5% | 45.8 | ***63*** | 93.2% | 64.9 |
| *27* | 37.5% | 46.8 | ***64*** | 93.7% | 65.3 |
| *28* | 41.9% | 47.9 | ***65*** | 94.0% | 65.6 |
| *29* | 46.0% | 49 | ***66*** | 94.3% | 65.8 |
| *30* | 49.6% | 49.9 | ***67*** | 94.6% | 66 |
| *31* | 53.1% | 50.8 | ***68*** | 94.8% | 66.3 |
| *32* | 56.5% | 51.6 | ***69*** | 95.2% | 66.6 |
| *33* | 59.6% | 52.4 | ***70*** | 95.4% | 66.9 |
| *34* | 62.5% | 53.2 | ***71*** | 95.6% | 67 |
| *35* | 65.3% | 53.9 | ***72*** | 95.8% | 67.2 |
| *36* | 67.9% | 54.6 | ***73*** | 96.0% | 67.5 |
| *37* | 70.1% | 55.3 | ***74*** | 96.3% | 67.8 |
| *38* | 72.0% | 55.8 | ***75*** | 96.6% | 68.2 |
| *39* | 73.7% | 56.3 | ***76*** | 96.7% | 68.4 |
| *40* | 75.5% | 56.9 | ***77*** | 96.9% | 68.7 |
| *41* | 77.1% | 57.4 | ***78*** | 97.1% | 69 |
| *42* | 78.2% | 57.8 | ***79*** | 97.3% | 69.2 |
| *43* | 79.6% | 58.3 | ***80**** | 97.3% | 69.3 |
| *44* | 80.8% | 58.7 | ***81*** | 97.4% | 69.4 |
| *45* | 81.8% | 59.1 | ***82*** | 97.6% | 69.8 |
| *46* | 82.7% | 59.4 | ***83*** | 97.8% | 70.2 |
| *47* | 83.6% | 59.8 | ***84*** | 98.0% | 70.5 |
| *48* | 84.8% | 60.3 | ***85*** | 98.1% | 70.7 |
| *49* | 85.6% | 60.6 | ***86*** | 98.2% | 71 |
| *50* | 86.1% | 60.9 | ***87*** | 98.4% | 71.5 |
| *51* | 86.7% | 61.1 | ***88*** | 98.5% | 71.8 |
| *52* | 87.2% | 61.4 | ***89*** | 98.7% | 72.2 |
| *53* | 87.7% | 61.6 | ***90*** | 98.8% | 72.5 |
| *54* | 88.3% | 61.9 | ***91*** | 98.9% | 73.1 |
| *55* | 89.0% | 62.3 | ***92*** | 99.1% | 73.8 |
| *56* | 89.6% | 62.6 | ***93*** | 99.3% | 74.4 |
| *57* | 90.1% | 62.9 | ***94*** | 99.4% | 74.9 |

**Combined Reactivity (Cont.)**

| *Raw Score* | *Percentile Rank* | *T-Score* | *Raw Score* | *Percentile Rank* | *T-Score* |
| --- | --- | --- | --- | --- | --- |
| *95* | 99.4% | 75.2 | ***101**** | 99.7% | 77 |
| *96* | 99.5% | 75.5 | ***102*** | 99.7% | 77.2 |
| *97* | 99.5% | 75.9 | ***103*** | 99.7% | 77.8 |
| *98* | 99.6% | 76.3 | ***104**** | 99.8% | 78.9 |
| *99* | 99.6% | 76.7 | ***105*** | 99.9% | 80.7 |
| *100** | 99.6% | 76.9 |  |  |  |

Study 2

*Physiological data collection prep.* First, participants were instructed to place Ag/AgCl wet liquid gel electrodes (7% chloride salt; Biopac EL503) in a Lead II configuration on their chest and abdomen. A diagram of the configuration and access to a private bathroom were provided. Research assistants visually checked electrode placement and helped with corrections as needed. Participants were seated and had two disposable Ag/AgCl electrodes (.5% chloride salt; Biopac EL507) placed by research assistants on the thenar and hypothenar eminences of the right palm for measurement of electrodermal activity (EDA). These electrodes were 27 mm x 36 mm, had a 11mm diameter contact area, and were prefilled with isotonic gel in a 1.5mm deep, 16mm diameter cavity. The participants’ outstretched arm was then placed to rest on a small table to the side of their chair.

ECG data were streamed using the BioNomadix wireless module (BN-RSPEC). The Biopac BN-PPGED amplifier was used to apply constant voltage (0.5 V) for EDA and measure current flow. Prior to recording, signal quality was visually inspected and electrodes were adjusted as needed. Lab temperature was maintained at approximately 21^o^C during testing.

*Associations between physiological variables and demographics*. VE and all HP, RSA, and SCR rate leg lift change scores were not associated with age, sex, or BMI (all p > .15). Absolute values for HP, RSA, or SCR rate at individual conditions (baseline, leg lifts, and recovery) did not differ by sex (all p > .20). Older age was associated with lower RSA (B = -0.08, SE = 0.03, t = -3.08, p = 0.003), but had no association with SCR rate or HP (all p > .15). Higher BMI was associated with lower RSA (B = -0.07, SE = 0.02, t = -2.84, p = 0.006) and lower HP (B = -5.92, SE = 2.00, t = -2.96, p = 0.004), but had no association with SCR rate (p > .10).

Table S8. Within-group contrasts for the plotted models in center row of Figure 2.

| Absolute Value Results |  |  |  |  |  |  |  |
| --- | --- | --- | --- | --- | --- | --- | --- |
| **Total Autonomic Symptoms (BPQ)** | **Contrast** | **Estimated Mean Difference** | **SE** | **df** | **t ratio** | **P-value** | **Effect Size** |
| Heart Period (ms) | | | | | | | |
| Low | Baseline - Leg Lift | 57.21 | 6.24 | 118 | 9.17 | <0.001 | 2.41 |
|  | Baseline - Recovery | 9.62 | 6.24 | 118 | 1.54 | 0.126 | 0.41 |
|  | Leg Lift - Recovery | -47.59 | 6.24 | 118 | -7.63 | <0.001 | -2.00 |
| Mid | Baseline - Leg Lift | 64.53 | 7.16 | 118 | 9.01 | <0.001 | 2.72 |
|  | Baseline - Recovery | 26.42 | 7.16 | 118 | 3.69 | <0.001 | 1.11 |
|  | Leg Lift - Recovery | -38.12 | 7.16 | 118 | -5.32 | <0.001 | -1.60 |
| High | Baseline - Leg Lift | 48.91 | 10.13 | 118 | 4.83 | <0.001 | 2.06 |
|  | Baseline - Recovery | 5.14 | 10.13 | 118 | 0.51 | 0.613 | 0.22 |
|  | Leg Lift - Recovery | -43.77 | 10.13 | 118 | -4.32 | <0.001 | -1.84 |
| RSA [ln(ms)^2] | | | | | | | |
| Low | Baseline - Leg Lift | 0.43 | 0.10 | 118 | 4.25 | <0.001 | 1.12 |
|  | Baseline - Recovery | 0.10 | 0.10 | 118 | 0.94 | 0.347 | 0.25 |
|  | Leg Lift - Recovery | -0.33 | 0.10 | 118 | -3.31 | 0.001 | -0.87 |
| Mid | Baseline - Leg Lift | 0.53 | 0.12 | 118 | 4.56 | <0.001 | 1.37 |
|  | Baseline - Recovery | 0.07 | 0.12 | 118 | 0.57 | 0.570 | 0.17 |
|  | Leg Lift - Recovery | -0.46 | 0.12 | 118 | -3.99 | <0.001 | -1.20 |
| High | Baseline - Leg Lift | 0.15 | 0.16 | 118 | 0.91 | 0.363 | 0.39 |
|  | Baseline - Recovery | -0.14 | 0.16 | 118 | -0.86 | 0.392 | -0.37 |
|  | Leg Lift - Recovery | -0.29 | 0.16 | 118 | -1.77 | 0.079 | -0.76 |
| SCR (Rate per Minute) | | | | | | | |
| Low | Baseline - Leg Lift |  |  |  |  | 0.012 | -0.60 |
|  | Baseline - Recovery |  |  |  |  | 0.166 | -0.33 |
|  | Leg Lift - Recovery |  |  |  |  | 0.016 | 0.58 |
| Mid | Baseline - Leg Lift |  |  |  |  | 0.009 | -0.71 |
|  | Baseline - Recovery |  |  |  |  | 0.007 | -0.73 |
|  | Leg Lift - Recovery |  |  |  |  | 0.952 | 0.02 |
| High | Baseline - Leg Lift |  |  |  |  | 0.281 | -0.60 |
|  | Baseline - Recovery |  |  |  |  | 0.529 | 0.33 |
|  | Leg Lift - Recovery |  |  |  |  | 0.106 | 0.87 |

Table S9. Within-group contrasts for the plotted models in bottom row of Figure 2.

| Change Score Results |  |  |  |  |  |  |  |
| --- | --- | --- | --- | --- | --- | --- | --- |
| **Total Autonomic Symptoms (BPQ)** | **Contrast** | **Estimated Mean Difference** | **SE** | **df** | **t ratio** | **P-value** | **Effect Size** |
| Heart Period Change (ms) | | | | | | | |
| Low | (Baseline - Leg Lift) - (Baseline - Recovery) | -47.59 | 7.55 | 59 | -6.30 | <0.001 | -1.66 |
| Mid | (Baseline - Leg Lift) - (Baseline - Recovery) | -38.12 | 8.67 | 59 | -4.40 | <0.001 | -1.33 |
| High | (Baseline - Leg Lift) - (Baseline - Recovery) | -43.77 | 12.26 | 59 | -3.57 | <0.001 | -1.52 |
| RSA Change [ln(ms)^2] | | | | | | | |
| Low | (Baseline - Leg Lift) - (Baseline - Recovery) | -0.33 | 0.11 | 59 | -3.14 | 0.003 | -0.82 |
| Mid | (Baseline - Leg Lift) - (Baseline - Recovery) | -0.46 | 0.12 | 59 | -3.79 | <0.001 | -1.14 |
| High | (Baseline - Leg Lift) - (Baseline - Recovery) | -0.29 | 0.17 | 59 | -1.68 | 0.098 | -0.72 |
| SCR Change (Rate per Minute) | | | | | | | |
| Low | (Baseline - Leg Lift) - (Baseline - Recovery) | 1.70 | 0.52 | 50 | 3.30 | 0.002 | 0.93 |
| Mid | (Baseline - Leg Lift) - (Baseline - Recovery) | 0.31 | 0.58 | 50 | 0.54 | 0.589 | 0.17 |
| High | (Baseline - Leg Lift) - (Baseline - Recovery) | 0.91 | 0.91 | 50 | 1.00 | 0.320 | 0.50 |

Table S9. Within-group contrasts for sub-diaphragmatic absolute value models.

| Absolute Value Results |  |  |  |  |  |  |  |
| --- | --- | --- | --- | --- | --- | --- | --- |
| **Sub-diaphragmatic symptoms** | **Contrast** | **Estimated Mean Difference** | **SE** | **df** | **t ratio** | **P-value** | **Effect Size** |
| Heart Period (ms) | | | | | | | |
| Low | Baseline - Leg Lift | 60.17 | 5.79 | 118 | 10.39 | <0.001 | 2.52 |
|  | Baseline - Recovery | 12.85 | 5.79 | 118 | 2.22 | 0.028 | 0.54 |
|  | Leg Lift - Recovery | -47.32 | 5.79 | 118 | -8.17 | <0.001 | -1.98 |
| Mid | Baseline - Leg Lift | 63.71 | 8.19 | 118 | 7.78 | <0.001 | 2.67 |
|  | Baseline - Recovery | 22.93 | 8.19 | 118 | 2.80 | 0.006 | 0.96 |
|  | Leg Lift - Recovery | -40.78 | 8.19 | 118 | -4.98 | <0.001 | -1.71 |
| High | Baseline - Leg Lift | 44.38 | 10.18 | 118 | 4.36 | <0.001 | 1.86 |
|  | Baseline - Recovery | 8.20 | 10.18 | 118 | 0.80 | 0.422 | 0.34 |
|  | Leg Lift - Recovery | -36.18 | 10.18 | 118 | -3.55 | <0.001 | -1.52 |
| RSA [ln(ms)^2] | | | | | | | |
| Low | Baseline - Leg Lift | 0.45 | 0.09 | 118 | 4.89 | <0.001 | 1.19 |
|  | Baseline - Recovery | 0.15 | 0.09 | 118 | 1.58 | 0.118 | 0.38 |
|  | Leg Lift - Recovery | -0.31 | 0.09 | 118 | -3.32 | 0.001 | -0.80 |
| Mid | Baseline - Leg Lift | 0.51 | 0.13 | 118 | 3.89 | <0.001 | 1.33 |
|  | Baseline - Recovery | -0.04 | 0.13 | 118 | -0.28 | 0.781 | -0.10 |
|  | Leg Lift - Recovery | -0.55 | 0.13 | 118 | -4.17 | <0.001 | -1.43 |
| High | Baseline - Leg Lift | 0.15 | 0.16 | 118 | 0.93 | 0.355 | 0.40 |
|  | Baseline - Recovery | -0.15 | 0.16 | 118 | -0.93 | 0.352 | -0.40 |
|  | Leg Lift - Recovery | -0.30 | 0.16 | 118 | -1.86 | 0.065 | -0.79 |
| SCR (Rate per Minute) | | | | | | | |
| Low | Baseline - Leg Lift |  |  |  |  | 0.011 | -0.56 |
|  | Baseline - Recovery |  |  |  |  | 0.140 | -0.33 |
|  | Leg Lift - Recovery |  |  |  |  | 0.010 | 0.57 |
| Mid | Baseline - Leg Lift |  |  |  |  | 0.021 | -0.77 |
|  | Baseline - Recovery |  |  |  |  | 0.025 | -0.74 |
|  | Leg Lift - Recovery |  |  |  |  | 1.000 | 0.01 |
| High | Baseline - Leg Lift |  |  |  |  | 0.151 | -0.64 |
|  | Baseline - Recovery |  |  |  |  | 0.834 | -0.11 |
|  | Leg Lift - Recovery |  |  |  |  | 0.554 | 0.29 |

Table S10. Within-group contrasts for sub-diaphragmatic change score models.

| Change Score Results |  |  |  |  |  |  |  |
| --- | --- | --- | --- | --- | --- | --- | --- |
| **Sub-diaphragmatic symptoms** | **Contrast** | **Estimated Mean Difference** | **SE** | **df** | **t ratio** | **P-value** | **Effect Size** |
| Heart Period Change (ms) | | | | | | | |
| Low | (Baseline - Leg Lift) - (Baseline - Recovery) | -47.32 | 6.97 | 59 | -6.79 | <0.001 | -1.65 |
| Mid | (Baseline - Leg Lift) - (Baseline - Recovery) | -40.78 | 9.85 | 59 | -4.14 | <0.001 | -1.42 |
| High | (Baseline - Leg Lift) - (Baseline - Recovery) | -36.18 | 12.25 | 59 | -2.95 | 0.005 | -1.26 |
| RSA Change [ln(ms)^2] | | | | | | | |
| Low | (Baseline - Leg Lift) - (Baseline - Recovery) | -0.31 | 0.10 | 59 | -3.16 | 0.002 | -0.77 |
| Mid | (Baseline - Leg Lift) - (Baseline - Recovery) | -0.55 | 0.14 | 59 | -3.97 | <0.001 | -1.36 |
| High | (Baseline - Leg Lift) - (Baseline - Recovery) | -0.30 | 0.17 | 59 | -1.77 | 0.081 | -0.76 |
| SCR Change (Rate per Minute) | | | | | | | |
| Low | (Baseline - Leg Lift) - (Baseline - Recovery) | 1.71 | 0.47 | 50 | 3.61 | <0.001 | 0.95 |
| Mid | (Baseline - Leg Lift) - (Baseline - Recovery) | 0.19 | 0.66 | 50 | 0.29 | 0.775 | 0.10 |
| High | (Baseline - Leg Lift) - (Baseline - Recovery) | 0.41 | 0.85 | 50 | 0.48 | 0.636 | 0.22 |

Table S11. Within-group contrasts for supra-diaphragmatic absolute value models.

| Absolute Value Results |  |  |  |  |  |  |  |
| --- | --- | --- | --- | --- | --- | --- | --- |
| **Supra-diaphragmatic symptoms** | **Contrast** | **Estimated Mean Difference** | **SE** | **df** | **t ratio** | **P-value** | **Effect Size** |
| Heart Period (ms) | | | | | | | |
| Low | Baseline - Leg Lift | 60.11 | 7.22 | 118 | 8.32 | <0.001 | 2.51 |
|  | Baseline - Recovery | 11.28 | 7.22 | 118 | 1.56 | 0.121 | 0.47 |
|  | Leg Lift - Recovery | -48.83 | 7.22 | 118 | -6.76 | <0.001 | -2.04 |
| Mid | Baseline - Leg Lift | 60.54 | 6.09 | 118 | 9.95 | <0.001 | 2.53 |
|  | Baseline - Recovery | 20.01 | 6.09 | 118 | 3.29 | 0.001 | 0.84 |
|  | Leg Lift - Recovery | -40.53 | 6.09 | 118 | -6.66 | <0.001 | -1.69 |
| High | Baseline - Leg Lift | 46.43 | 11.29 | 118 | 4.11 | <0.001 | 1.94 |
|  | Baseline - Recovery | 5.37 | 11.29 | 118 | 0.48 | 0.635 | 0.22 |
|  | Leg Lift - Recovery | -41.06 | 11.29 | 118 | -3.64 | <0.001 | -1.71 |
| RSA [ln(ms)^2] | | | | | | | |
| Low | Baseline - Leg Lift | 0.58 | 0.12 | 118 | 4.97 | <0.001 | 1.50 |
|  | Baseline - Recovery | 0.05 | 0.12 | 118 | 0.46 | 0.643 | 0.14 |
|  | Leg Lift - Recovery | -0.52 | 0.12 | 118 | -4.51 | <0.001 | -1.36 |
| Mid | Baseline - Leg Lift | 0.36 | 0.10 | 118 | 3.66 | <0.001 | 0.93 |
|  | Baseline - Recovery | 0.08 | 0.10 | 118 | 0.84 | 0.404 | 0.21 |
|  | Leg Lift - Recovery | -0.28 | 0.10 | 118 | -2.82 | 0.006 | -0.72 |
| High | Baseline - Leg Lift | 0.23 | 0.18 | 118 | 1.24 | 0.216 | 0.59 |
|  | Baseline - Recovery | -0.12 | 0.18 | 118 | -0.64 | 0.522 | -0.30 |
|  | Leg Lift - Recovery | -0.34 | 0.18 | 118 | -1.89 | 0.062 | -0.89 |
| SCR (Rate per Minute) | | | | | | | |
| Low | Baseline - Leg Lift |  |  |  |  | 0.011 | -0.71 |
|  | Baseline - Recovery |  |  |  |  | 0.115 | -0.46 |
|  | Leg Lift - Recovery |  |  |  |  | 0.127 | 0.42 |
| Mid | Baseline - Leg Lift |  |  |  |  | 0.018 | -0.54 |
|  | Baseline - Recovery |  |  |  |  | 0.051 | -0.44 |
|  | Leg Lift - Recovery |  |  |  |  | 0.090 | 0.39 |
| High | Baseline - Leg Lift |  |  |  |  | 0.201 | -0.80 |
|  | Baseline - Recovery |  |  |  |  | 0.281 | -0.60 |
|  | Leg Lift - Recovery |  |  |  |  | 0.361 | 0.60 |

Table S12. Within-group contrasts for supra-diaphragmatic change score models.

| Change Score Results |  |  |  |  |  |  |  |
| --- | --- | --- | --- | --- | --- | --- | --- |
| **Supra-diaphragmatic symptoms** | **Contrast** | **Estimated Mean Difference** | **SE** | **df** | **t ratio** | **P-value** | **Effect Size** |
| Heart Period Change (ms) | | | | | | | |
| Low | (Baseline - Leg Lift) - (Baseline - Recovery) | -48.83 | 8.67 | 59 | -5.63 | <0.001 | -1.70 |
| Mid | (Baseline - Leg Lift) - (Baseline - Recovery) | -40.53 | 7.31 | 59 | -5.55 | <0.001 | -1.41 |
| High | (Baseline - Leg Lift) - (Baseline - Recovery) | -41.06 | 13.56 | 59 | -3.03 | 0.004 | -1.43 |
| RSA Change [ln(ms)^2] | | | | | | | |
| Low | (Baseline - Leg Lift) - (Baseline - Recovery) | -0.52 | 0.12 | 59 | -4.32 | <0.001 | -1.30 |
| Mid | (Baseline - Leg Lift) - (Baseline - Recovery) | -0.28 | 0.10 | 59 | -2.70 | 0.009 | -0.69 |
| High | (Baseline - Leg Lift) - (Baseline - Recovery) | -0.34 | 0.19 | 59 | -1.81 | 0.076 | -0.85 |
| SCR Change (Rate per Minute) | | | | | | | |
| Low | (Baseline - Leg Lift) - (Baseline - Recovery) | 1.31 | 0.61 | 50 | 2.16 | 0.036 | 0.70 |
| Mid | (Baseline - Leg Lift) - (Baseline - Recovery) | 1.07 | 0.51 | 50 | 2.11 | 0.040 | 0.58 |
| High | (Baseline - Leg Lift) - (Baseline - Recovery) | 0.33 | 1.00 | 50 | 0.33 | 0.744 | 0.18 |

Figure S4. Sub-diaphragmatic symptom model results

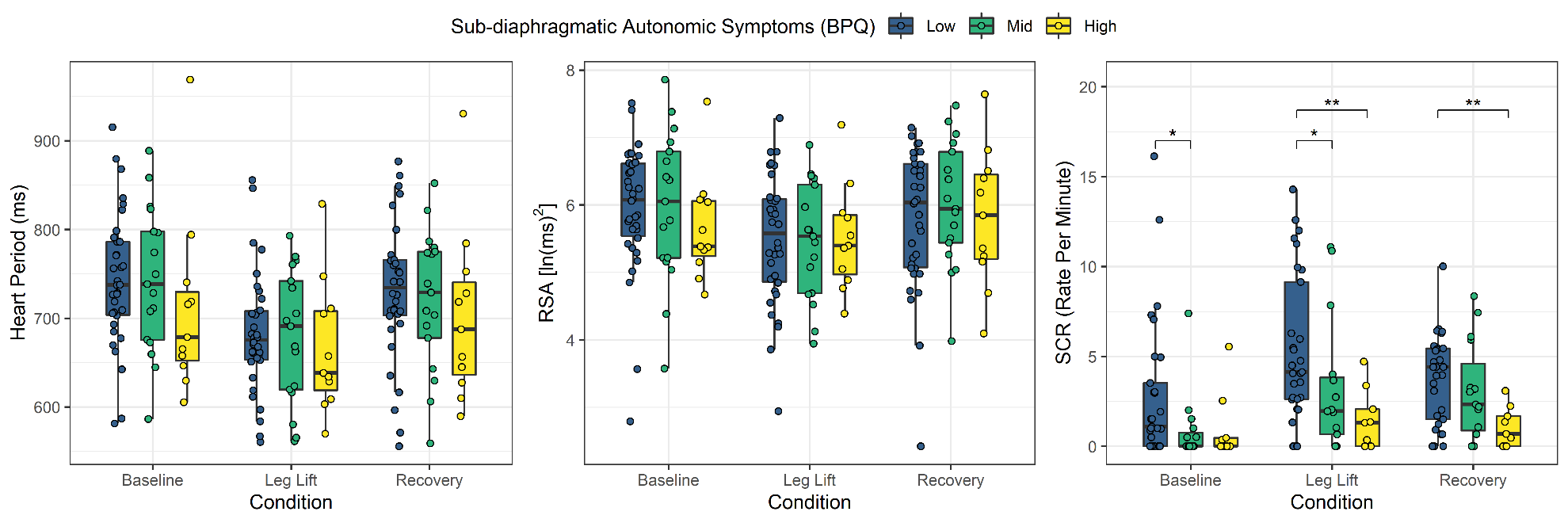

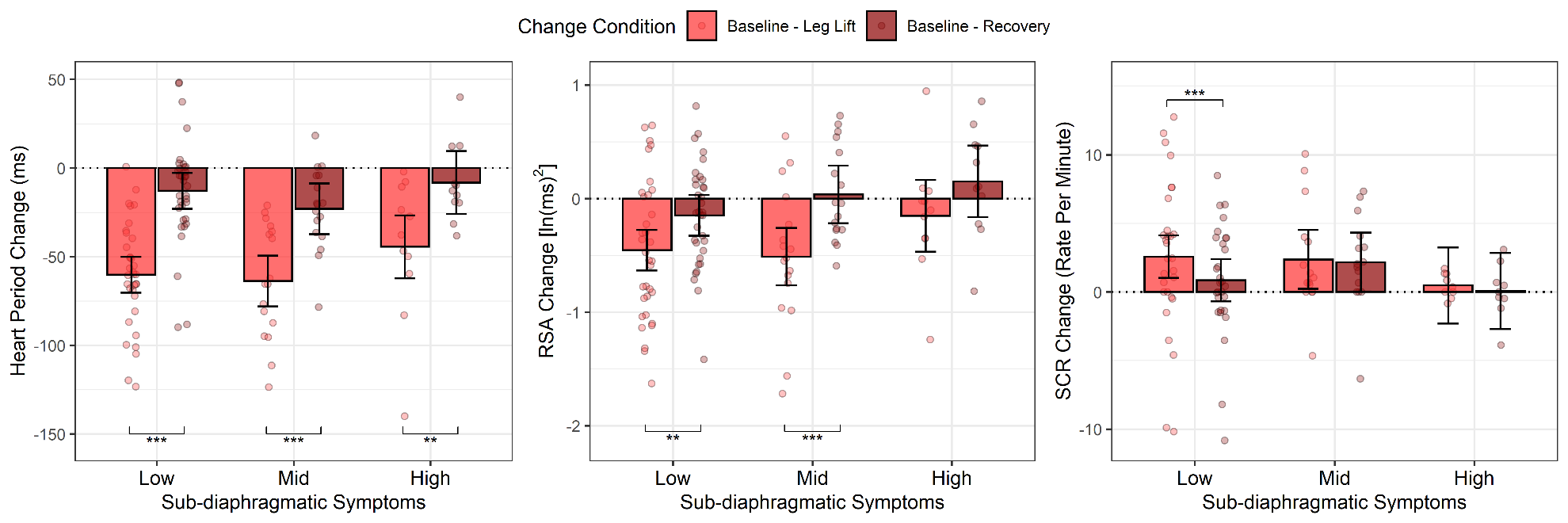

Figure S5. Supra-diaphragmatic symptom model results

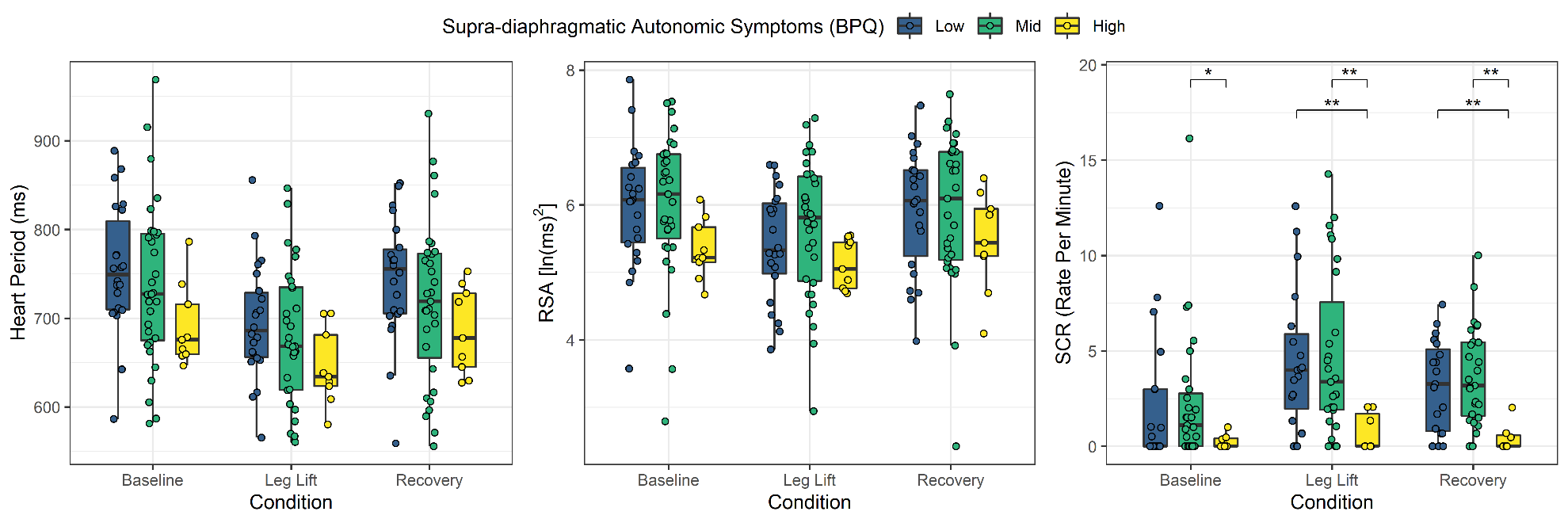

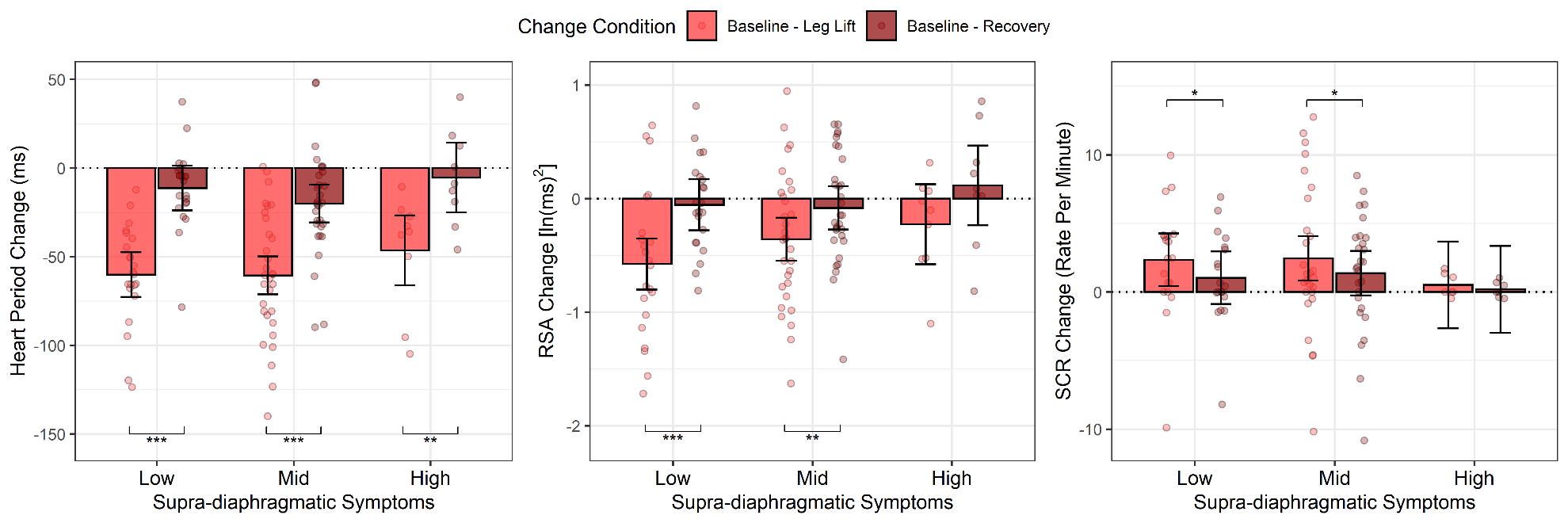
